# Interpretable Machine Learning Leverages Proteomics to Improve Cardiovascular Disease Risk Prediction and Biomarker Identification

**DOI:** 10.1101/2024.01.12.24301213

**Authors:** Héctor Climente-González, Min Oh, Urszula Chajewska, Roya Hosseini, Sudipto Mukherjee, Wei Gan, Matthew Traylor, Sile Hu, Ghazaleh Fatemifar, Paul Pangilinan Del Villar, Erik Vernet, Nils Koelling, Liang Du, Robin Abraham, Chuan Li, Joanna M. M. Howson

**Author notes:** Equal contribution (Héctor Climente-González, Min Oh). Equal contribution (Roya Hosseini, Sudipto Mukherjee). Corresponding authors: Chuan Li; Joanna M. M. Howson.

## Abstract

Cardiovascular diseases (CVD), primarily coronary heart disease and stroke, rank amongst the leading causes of long-term disability and mortality. Providing accurate disease risk predictions and identifying genes associated with CVD are crucial for prevention, early intervention, and the development of novel medications.

The recent availability of UK Biobank Proteomics data enables the investigation of the blood proteome and its association with a wide variety of diseases. We employed the Explainable Boosting Machine (EBM), an interpretable machine learning model, for CVD risk prediction. The EBM model using proteomics outperforms traditional clinical models with an AUROC of 0.767 and an AUPRC of 0.2405. Adding clinical features further improves the AUROC to 0.785 and the AUPRC to 0.2835. Our models demonstrate consistent performance across sexes and ethnicities.

While most prior studies using proteomics data for disease prediction have primarily focused on maximizing the accuracy at the population level, our model provides additional enriched insights into individualized disease risk predictions and in-depth biological insights into biomarkers. Our analysis also uncovers nonlinear risks linked to varying feature values. We further corroborate our findings using statistical approaches and evidence from the literature.

In conclusion, we present a highly accurate and explanatory framework for proteomics data analysis, offering comprehensive and in-depth molecular and clinical insights. Our findings support future approaches that prioritize individualized disease risk prediction and the identification of target genes for drug development.

## Introduction

Cardiovascular disease (CVD), primarily coronary heart disease and stroke, collectively rank among the leading causes of death worldwide. The primary pathology underlying these diseases is atherosclerosis, which is characterized by the build-up of plaque in major arteries. CVD impacts people in both developed and developing countries, encompassing individuals from diverse ethnic backgrounds and has an increasing impact on women and younger individuals (Libby, 2021). Current treatment approaches include lifestyle adjustments, regular medical monitoring, and pharmaceutical interventions targeting major risk factors such as elevated cholesterol (statins) and blood pressure (Charo & Taub, 2011; Chen, et al., 2022). To enhance existing treatments, identify individuals at risk of disease, and improve preventative measures, current approaches may be augmented by accurate personal disease risk predictions. Traditional clinical predictors, such as blood pressure, BMI, cholesterol levels, and medical and family history, have been employed to estimate individual disease risk, but their accuracy remains limited (Singh, Pilkerton, Shrader, & Frisbee, 2018; Huang, et al., 2022). Therefore, there is a need for developing more precise predictive methods incorporating state-of-the-art omics technologies such as proteomics.

The UK Biobank recruited over 500,000 participants to gather comprehensive baseline data and long-term follow-up of health outcomes (Bycroft, et al., 2018). To promote an in-depth understanding of disease biology and accelerate drug development, proteomics data were collected from over 54,000 participants (Sun, et al., 2023). The circulating concentration of 3,072 plasma proteins was quantified using Proximity Extension Assay with the Olink Explore Platform. This proteomics data can be utilized for drug target and biomarker discovery, to improve disease understanding, and to inform patient stratification as well as disease prediction. Here, we focus on building machine learning models for two primary objectives: predicting disease risk and identifying genes associated with it.

Previous studies have used proteomics to predict cardiovascular events (Helgason, et al., 2023), type II diabetes (Gadd, et al., 2023), and chronic kidney disease (Avram, 2023). Incorporating additional information, such as clinical (Williams, et al., 2022), lipidomics (Nurmohamed, et al., 2023), and metabolomics data (Nightingale Health Biobank Collaborative Group, et al., 2023) has been demonstrated to further enhance the predictive capabilities of disease risk. Apart from predicting disease risk, some machine learning models also excel in extracting in-depth insights of feature importance and individualized risk predictions from high-dimensional and complex datasets. For instance, Schuermans et al. recently applied LASSO regression to predict common cardiac diseases in the initial proteomics data released by the UK Biobank, identifying 820 potential protein-disease associations (Schuermans, et al., 2023). However, such linear models may not fully capture non-linear relationships between predictors and outcomes.

Explainable Boosting Machines (EBMs) are interpretable and non-linear machine learning models that belong to the family of Generalized Additive Models (GAMs) (Hastie & Tibshirani, 1987). These models involve a response variable that depends linearly on shape functions, which are unknown smooth functions of predictor variables. EBMs use bagged ensembles of boosted depth-restricted trees to represent these shape functions (Lou, Caruana, & Gehrke, 2013). In simpler terms, EBMs use a combination of simple, single-feature models to accurately represent how each of the features (e.g., age or plasma level of a protein) predict an outcome (disease risk) by using a technique called boosting. In boosting models, simple models are created sequentially, with each new model trying to correct the errors made by the previous ones. This process helps to create a robust final model that can provide accurate and reliable predictions. When the final model is built, the per-feature models get combined into the shape functions, which provide valuable insights into the risk profile for subranges of features’ values. EBM models offer both local and global explanations. Local explanations predict the features (e.g., molecular or clinical factors) that are important for each participant. For instance, the local explanation for a participant’s prediction might show that a particular protein is an important predictor of disease risk, based on its expression level in that participant. On the other hand, global explanations focus on the importance of features across a spectrum of values. For instance, the global explanation for a specific protein would show how risk changes across the different expression levels observed in the whole cohort. For comparison, we also considered predictive models using other gradient boosting approaches popular in the literature in addition to EBM. Gradient boosted trees are well-established strong baselines for tabular data (Grinsztajn, Oyallon, & Varoquaux, 2022). Moreover, they are robust to uninformative features and outperform other methods on skewed data distributions. Coupled with SHapley Additive exPlanation (SHAP) values (Lundberg & Lee, 2017), these classifiers provide a way to explain the global and local feature importance as well.

In this study, we showcase not only the high predictive power of our machine learning models for CVD risk but also their explainability. Specifically, we apply the EBM, an explainable machine learning framework, for predicting CVD risk. Our analysis reveals insights into three aspects of explainability: (1) features with high predictive power; (2) participant-specific risk factors; and (3) distinct risks associated with varying feature values. We further substantiate our findings through statistical approaches and reviewing existing literature.

## Results

### Model accuracy of cardiovascular disease risk prediction

Using data from the UK Biobank participants with proteomics measurements, we aimed to predict the risk of a primary cardiovascular disease (CVD) event occurring within 10 years of the initial assessment center visit, in which blood samples were collected (see Methods Section on UKBB dataset for the definition of CVD). Supplementary material also includes results on 3 and 5-year risk predictions. Proteomic and clinical features were used to train the models. Specifically, after quality control exclusions (see Methods), plasma levels of 2,941 proteins quantified in 46,009 participants were included in our models (**Fig. 1A**).

**Figure 1.**
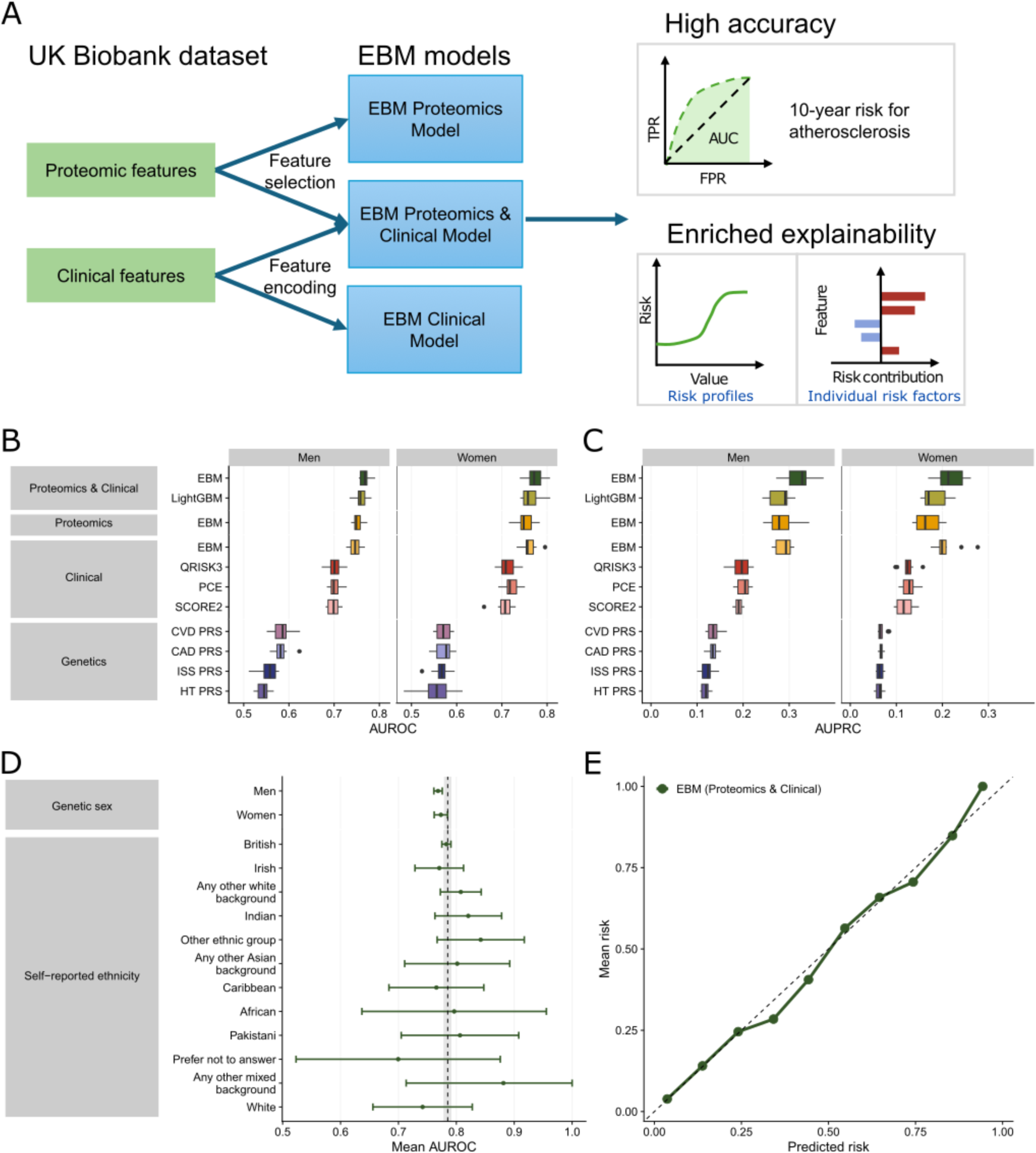
Experiment overview and high performance of Explainable Boosting Machine (EBM) models. **(A)** Schematic overview of the study design. This study focuses on using the UK Biobank proteomics and clinical data to predict target disease risk for cardiovascular disease (CVD) and identify target proteins using advanced machine learning models. Detailed insights were revealed for the risk profiles for genes and individual risk factors for patients. **(B-C)** EBM models significantly outperform the other models at predicting the 10-year risk of CVD. **(B)** Area under the receiver operating characteristic (AUROC) and **(C)** area under the precision-recall curve (AUPRC) are displayed for the polygenic risk scores (PRS), the clinical scores, and machine learning approaches trained on different sets of variables. The PRSs are for cardiovascular disease (CVD), coronary artery disease (CAD), hypertension (HT) and ischaemic stroke (ISS). The machine learning approaches are EBM and LightGBM. **(D)** Mean AUROC across the two genetic sexes and the self-reported ethnicities with at least 5 cases. The bars represent the 95% confidence interval. The dashed line represents the AUROC over the whole cohort, and the shaded area is its 95% confidence interval. **(E)** The predicted risk aligns well to the observed risk in the evaluation set.

Our proteomics-only model achieved an area under the receiver operating characteristic curve (AUROC) of 0.767 and an area under the precision-recall curve (AUPRC) of 0.241, outperforming three traditional CVD risk prediction models (PCE, QRISK3, and SCORE2), that rely on clinical features, and four relevant polygenic risk scores (PRSs) (**Fig. 1B-C**, **Table 1**). Our results hold for events in shorter timeframes (**Supplementary Figure 1**). We measured the improvement brought over by our models via the net reclassification improvement (NRI) and the integrated discrimination improvement (IDI) metrics. The model trained on proteomics data displayed significantly better performance than the best performing traditional score (QRISK3) for both metrics (**Table S1**).

**Table 1.**
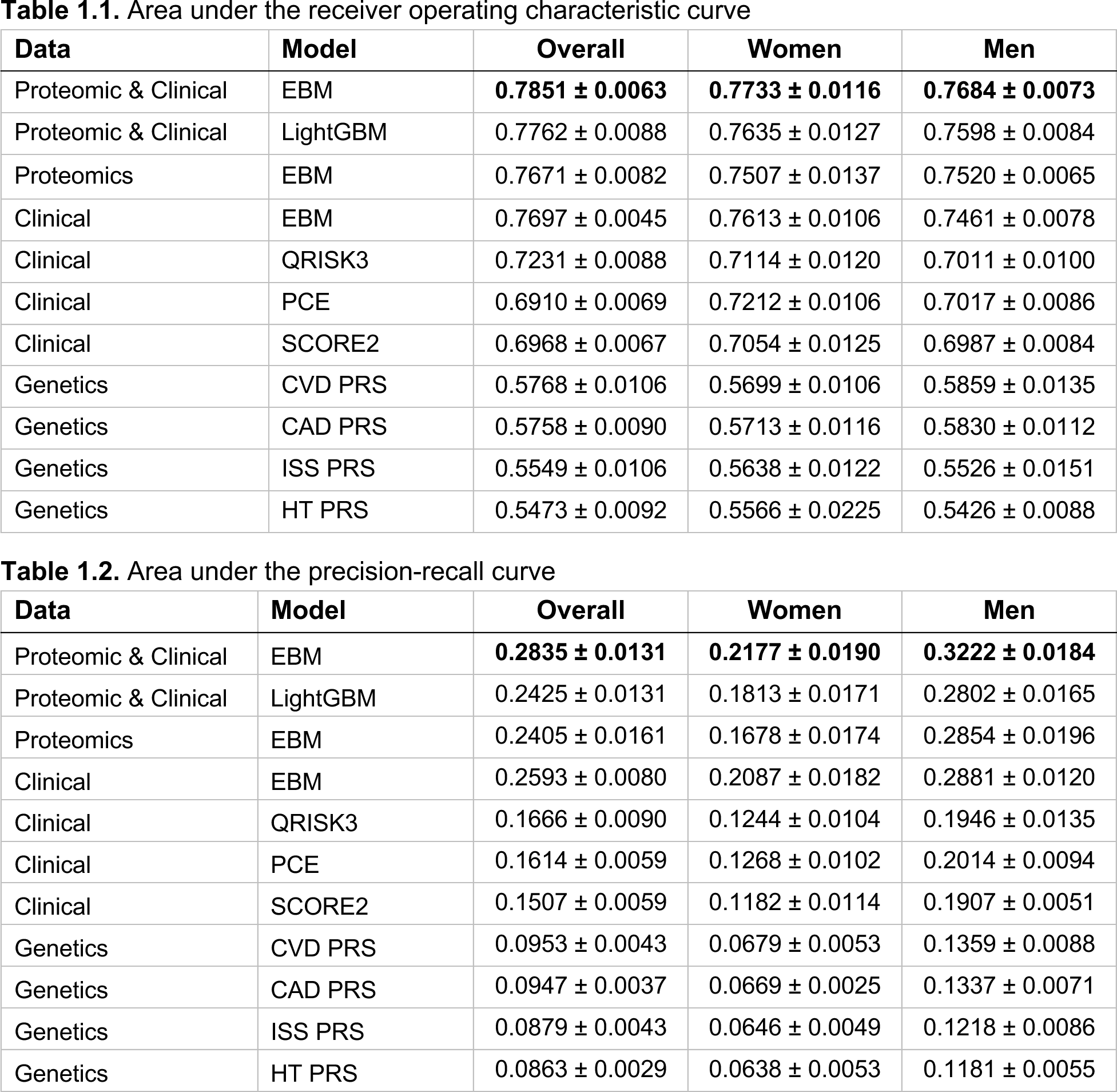
Mean performance and 95% confidence interval of the models shown in Figure 1B-C. The best performing model in each column is highlighted in bold.

To enhance predictive accuracy, we trained EBM models on both clinical and proteomics features (**Fig. 1B-C**), which increased the AUROC to 0.785 and the AUPRC to 0.284 (**Table 1**). The EBM Proteomics & Clinical model, trained on both clinical and proteomics data, outperformed both the EBM Proteomics model (ΔAUROC = 0.018, ΔAUPRC = 0.043), which was trained solely on proteomics features, and the EBM Clinical model (ΔAUROC = 0.0154, ΔAUPRC = 0.024), which focuses solely on clinical features. The EBM model also outperformed the LightGBM model, a state-of-the-art machine learning model for tabular data (ΔAUROC = 0.009, ΔAUPRC = 0.041). When examining its predictive performance at shorter time horizons, the EBM models remained competitive compared with the LightGBM model (**Supplementary** Figure 1).

### Model fairness and calibration

The EBM Proteomics & Clinical model shows consistent performance across genetic sexes and self-reported ethnicities (**Fig. 1D**). Crucially, it remains competitive across ethnicities, being the best performing model in most of them (**Supplementary Figure 2**). We evaluated whether our model remains predictive among the participants taking statins, commonly used to treat elevated LDL cholesterol (**Supplementary Figure 3**). The AUROC is 0.786 for people not taking statins, and 0.685 for people taking statins. This gap in model performance might be explained by the small number of participants that were both taking statins and had a primary CVD event: among 2,967 participants taking statins, only 318 participants (10.7%) had an event by year 10. Moreover, the performances across subpopulations with different income, blood pressure and LDL levels were largely consistent, all exceeding the current state-of-the-art clinical model (**Supplementary Figure 3**).

The EBM Proteomics & Clinical model showed similar performance at predicting specific acute complications like ischemic stroke and myocardial infarction, as well as diagnosis of coronary artery disease (CAD), with AUROC scores varying from 0.785 for CAD to 0.802 for stroke (**Supplementary Figure 4, Table S6**). In addition, for each subtype, the performance was comparable between sexes (**Supplementary Figure 4, Table S6**). The largest gap occurred for infarction, with an AUROC for women of 0.787 and for men of 0.767.

We also assessed the calibration of the EBM Proteomics & Clinical model by examining whether the predicted risk probability matches with the actual disease risk. This step is crucial in ensuring that the model produces reliable predictions with clinical relevance, i.e., the model score corresponds to the primary event probability of CVD. Overall, our model is well calibrated (**Fig. 1E**).

### Local and global explanations on feature importance

Besides their superior performance, the EBM models are highly interpretable, providing both local and global measures of feature importance. Local explanations quantify the contribution of different features to an individual participant’s predicted CVD risk. EBM can produce risk factors for each individual participant (**Fig. 2A-C**), which might vary considerably. For example, participants with similar predicted risks can have distinct sets of predictive proteomic features (**Fig. 2A-B**), which are distinct from a low-risk individual (**Fig. 2C**). Uncovering such individualized risk factors will aid in better understanding the underlying etiology and has potential to influence disease management. Global explanation refers to assessing the overall feature importance of each proteomic feature in the population. Notably, 5 out of the 10 most important predictors are known biomarkers of cardiovascular health (**Fig. 2D**): NT-proBNP (Cao, Jia, & Zhu, 2019), NPPB (Cao, Jia, & Zhu, 2019), PLA2G7 (Lp-PLA2) (Thompson, et al., 2010), MMP12 (Traylor, et al., 2014; Sun, et al., 2018) and GDF15 (Adela & Banerjee, 2015; Wang, et al., 2021; Wollert, Kempf, & Wallentin, 2017). The proteins with the largest feature importance were largely consistent across different time frames (**Supplementary Figure 5**). Notably, the top features revealed by EBM models were largely consistent with the LightGBM model represented by SHAP values (**Supplementary Figure 6**).

**Figure 2.**
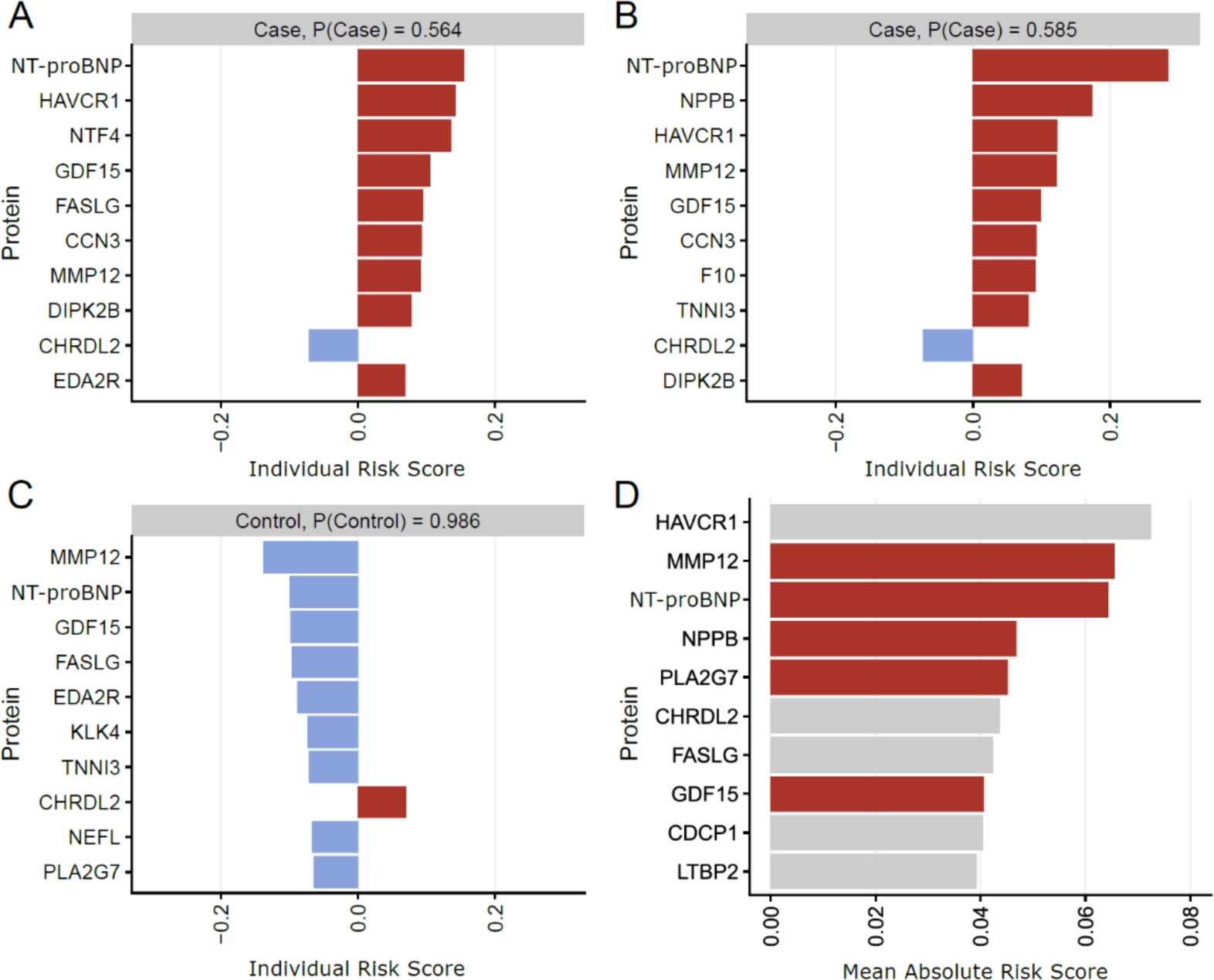
EBM models provide local and global explanations for feature importance. **(A-C)** Local explanations quantify the contribution of different features to an individual participant’s predicted CVD risk. The model quantifies the contribution of each protein to CVD risk for every participant. Red bars represent increased risks based on the plasma protein expression level, while blue bars denote reduced disease risks according to the expression level. Note participants in **(A)** and **(B)** show similar risk but different contributing proteins, while participant in **(C)** shows low disease risk. The intercept, with a value of −3.04, is not shown. **(D)** Global explanations aggregate the contribution of features across the cohort. The graph displays the top 10 contributing proteins in the EBM model. Known CVD markers were highlighted in red.

EBM models allow to study the risk profile for each feature across its full value range (**Fig. 3**). In many traditional machine learning models, feature importance is represented by a single value, which is not ideal for interpretability due to two main shortcomings. Firstly, a single value of feature importance tends to conflate the prevalence of an effect with the effect’s magnitude. That is, a protein with a very high risk restricted to a small number of participants would appear indistinguishable from that of a protein with a slightly elevated risk over many participants. For example, FASLG and GDF15 showed similar global feature importance value, but their risk profiles differ drastically (**Fig. 3A-B**). Similarly, two clinical features with similar feature importance could have different feature distributions and risk profiles in the EBM Clinical model (**Fig. 3C-D**). Secondly, traditional machine learning models disregard the dependence of risk on feature subranges. While a higher CVD risk is linked with the higher levels of gene GDF15, the risk does not increase linearly, instead plateauing at a very high level (**Supplementary Figure 7**). Our model offers detailed insights into the predicted disease risks associated with various circulating levels for each biomarker protein, contributing valuable insights into the disease mechanisms.

**Figure 3.**
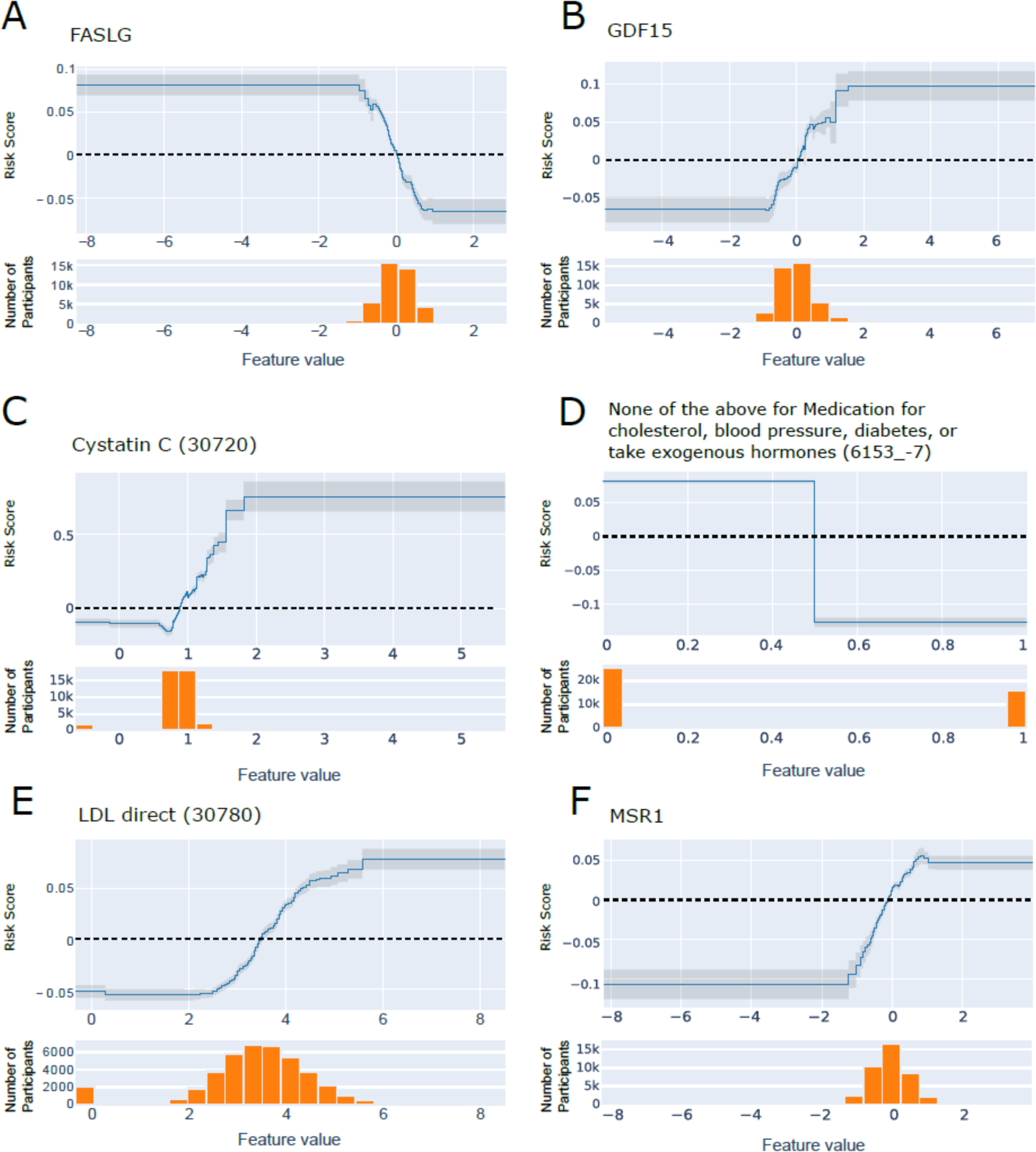
Risk profiles for selected top features. **(A)** FASLG and **(B)** GDF15 have similar feature importance in the EBM Proteomics & Clinical model, but the risk profiles differ drastically. Two clinical features **(C)** Cystatin C (field 30720) and **(D)** Medication history (field 6153, value −7) in the EBM Clinical model with similar risk profiles show drastically different risk profiles and feature distributions. **(C)** A small number of participants with high Cystatin C levels show greatly increased CVD risk. **(D)** Participants who selected “none of the above” when filling out medication for cholesterol, blood pressure, diabetes, or take exogenous hormone showed reduced risk of CVD. This is a weak effect affecting a large number of individuals. Risk profiles for **(E)** HDL (field 30780) and **(F)** MSR1 show high consistency. The top panels of each subfigure display the risk score on the logarithmic scale (y-axis) for different feature values (x-axis), with gray shaded regions representing the standard deviation for estimations. The dotted line corresponds to the population mean risk for the given feature. Positive values indicate increased risk; negative values indicate decreased risk. The bottom panels show histograms of the feature values across all participants.

Studying the EBM Proteomics & Clinical model allows to understand the contribution of clinical features to CVD risk prediction in a proteomics context. Among the 20 most important features in the model (**Table S2**) we find the clinical variables related to medications, age, sex, family history of heart disease and LDL levels. Notably, when features are correlated, the importance might get split across them. For instance, NT-proBNP shows reduced importance after incorporating clinical features (**Supplementary Figure 8**).

Furthermore, as with the protein levels, EBM models allowed us to study how risk changes across the feature ranges. For instance, it recapitulated how elevated LDL levels are predictive of a higher risk of CVD (**Fig. 3E**), while higher HDL levels are predictive of a lower risk of CVD (**Supplementary Figure 9A**) (The Emerging Risk Factors Collaboration, 2009). Similarly, we also recapitulated risk profiles of plasma proteins involved in cholesterol homeostasis. MSR1 (ranked 18 in the EBM Proteomics model) is responsible for mediating the endocytosis of modified LDLs and the uptake of modified lipoprotein (Sheng, Ji, & Zhang, 2022). In line with previous studies, elevated MSR1 levels were associated with an increased atherosclerosis risk (**Fig. 3F**) (Gudgeon, Marin-Rubio, & Trost, 2022). PCSK9 (ranked 19 in the EBM Proteomics model), which inhibits the clearance of plasma LDL by downregulating the LDL-C receptor, has been identified as a contributing factor to atherosclerosis risk when overexpressed (**Supplementary Figure 9B**) (Ma, Hou, & Liu, 2023).

### Pathway analysis and feature importance comparisons

To better understand the contributions from different molecular pathways, we grouped proteomic features into KEGG pathways involved in atherosclerosis (including KEGG: map05417 and related pathways; 13 in total) and calculated the aggregated contribution of each pathway (**Fig. 4A**). PI3K-Akt signaling, reflecting apoptosis and cell-cycle contributions to atherosclerosis, and cholesterol metabolism were the two pathways with the highest contributions.

**Figure 4.**
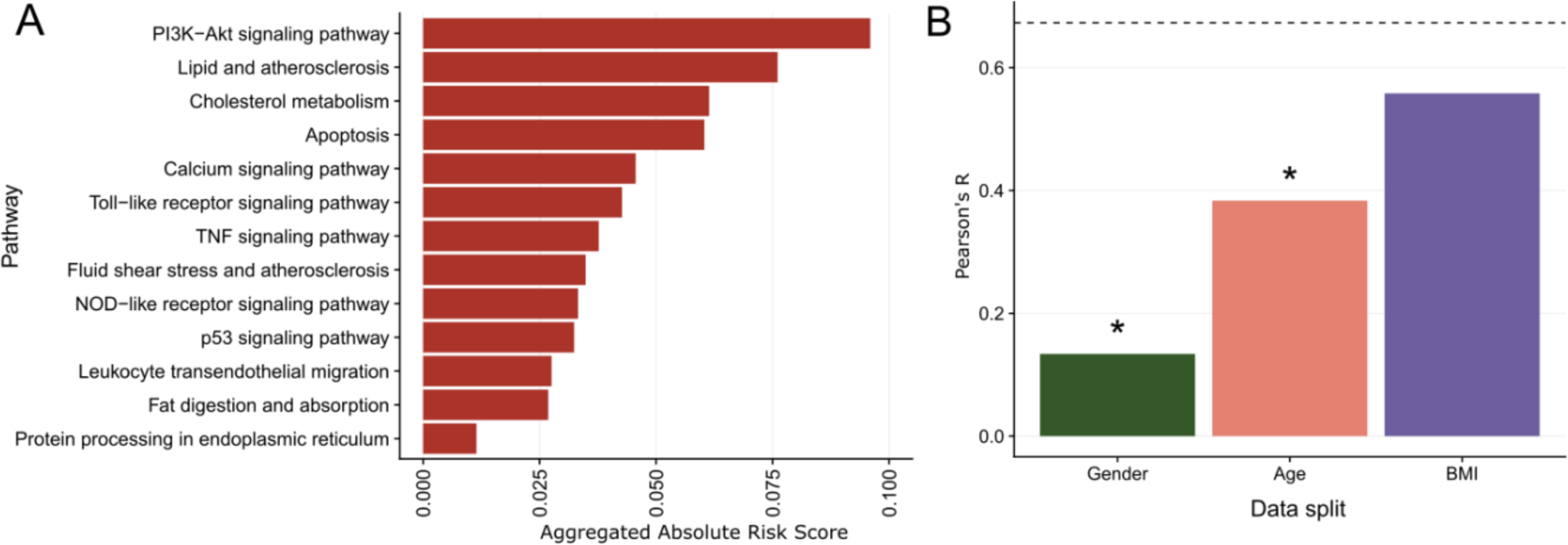
Aggregated pathway contribution and feature importance differences across subpopulations. **(A)** The aggregated contribution from top pathways predictive of CVD risk. **(B)** Models built based on a split by gender or age showed significantly lower linear correlations of feature importance between the subgroup models compared with random splits shown as dotted line. In contrast, no significant differences were observed for models based on BMI splits.

While the overall model performances are similar across genetic sexes, age groups and BMI profiles, we observe differences in predictive features in models trained separately on these subgroups. Only 2 and 3 of the top 10 features are shared between age and sex splits, respectively, while 5 features are shared between the high and low BMI splits (**Supplementary Figure 10**).

To verify that these differences are not due to stochastic noise, we randomly divided the participant population into two equal-sized groups and reran the experiment. Seven of the top 10 features are shared across the two groups, with correlated feature importance for the top 100 features (*R* = 0.67, **Supplementary Figure 11**). In contrast, we observe significant differences in feature importance for models built for different sexes (*R* = 0.13, *P* = 2e-9 compared with correlation between the random splits) and age groups (*R* = 0.38, *P* = 3e-4), suggesting that the underlying mechanisms might vary among these groups. However, no significant differences are found between the low and high BMI groups compared to the random split (*R* = 0.58, *P* = 0.10; **Fig. 4B, Supplementary Figure 11**).

## Discussion

In this study, we employed the EBM model to predict 10-year CVD risk using the UK Biobank proteomic data, generating models with high predictive power and explainability. By jointly considering the clinical and proteomic features, our model offered a comprehensive and coherent understanding of disease mechanisms, suggesting candidates for further clinical examination. The EBM Proteomics & Clinical model performance surpassed traditional models for CVD risk prediction like QRISK3 (ΔAUROC = 0.062, ΔAUPRC = 0.117) and the competing machine learning model, LightGBM (ΔAUROC = 0.009, ΔAUPRC = 0.041). The improvement in the AUPRC is particularly significant, given that CVD only affected 7.1% of our studied cohort. The EBM Clinical model and the EBM Proteomics model demonstrate similar performance (ΔAUROC = 0.003, ΔAUPRC = 0.018). However, a combination of both Proteomics and Clinical features results in further improvements on performance (ΔAUROC = 0.015, ΔAUPRC = 0.024). This underscores the consistency between Clinical and Proteomics features, and emphasizes that each offers unique information not provided by the other.

Our approach enables individualized risk prediction and generates enriched risk profiles for predictive features. As shown earlier, two participants exhibiting similar risks of CVD might have different features contributing to that prediction (**Fig. 2A-B**). By incorporating feature range-associated disease risk, our model can offer a comprehensive and holistic understanding of the prediction for each individual with a high level of interpretability. Furthermore, our approach also shows comparatively good performance at predicting CVD in shorter timeframes. This might be key to identify patients at high immediate risk.

Many of the proteins featured in this study have previously been reported as biomarkers for CVD. MMP12 (matrix metalloproteinase-12) levels have previously been shown to predict CVD (Goncalves, et al., 2015), as well as being directly involved in atherosclerosis pathogenesis (Newby, 2016), including from genetic studies (Traylor, et al., 2014; Sun, et al., 2018). NT-proBNP (**Supplementary** Figure 7A) and NPPB (**Supplementary** Figure 7B) exhibit similar risk profiles, as they are both derived from different fragments of the pro-B-type natriuretic peptide precursor. Each participate in the natriuretic peptide system, a vital pathway for regulating blood pressure and fluid balance, and are used in diagnosing cardiac dysfunction (Cao, Jia, & Zhu, 2019). PLA2G7, also known as Lp-PLA(2), is an inflammatory enzyme expressed in atherosclerotic plaques, and is a well-established marker of coronary heart disease (Thompson, et al., 2010). GDF15 is another prognostic marker for heart disease (Adela & Banerjee, 2015; Wollert, Kempf, & Wallentin, 2017; Wang, et al., 2021). Its expression level is increased by metformin, the primary treatment for diabetes, which aligns with diabetes being a risk factor for atherosclerosis. Furthermore, we also demonstrated the predictive ability of genes that were not previously used as CVD markers. For example, HAVCR1 was previously used as a biomarker for kidney injury (Han, Bailly, Abichandani, Thadhani, & Bonventre, 2002).

However, our approach has some limitations. First, most study participants were of European origin. Although we show consistent performance across ancestries, our model has limited generalizability to all patients. Further model development in non-European cohorts is necessary. Second, while certain features may be highly predictive, they do not necessarily imply a causal relationship. As such, further validation of our findings through experimental approaches and independent cohorts is essential to confirm causal relationships. Mendelian randomization and reviewing existing literature can be employed to further support or refute the causal relationships. Last, the proteomics panel contains a non-random subset of 2,941 proteins. A more extensive panel might lead to better models and the discovery of novel disease-protein associations.

Our findings underscore the importance and feasibility of individualized risk prediction and information-rich analysis of feature importance. The high accuracy of our proteomics model supports its potential for future clinical application in disease risk prediction using standalone proteomics blood tests. Additionally, our explanatory modeling framework can be readily adapted for predicting other diseases and phenotypes, providing valuable insights to facilitate drug development and individualized risk assessments.

## Methods

### UK Biobank Dataset

The UK Biobank is a large-scale prospective study designed to investigate the impact of biological and environmental factors on human health. It enrolled ∼500,000 participants aged 40-69 years old between 2006 and 2010 in England, Scotland, and Wales. In this study, we focused on the 54,181 participants that were included in the UK Biobank Pharma Proteomics Project (UKB-PPP) and had maintained their consent up until April 20, 2023. The UKB-PPP participants were representative of the entire UK Biobank cohort (Sun, et al., 2023). This research was conducted under the approved applications numbered 53639 and 65851.

Each sample was characterized by a rich set of 2,941 protein features, derived from two major releases: 1,472 features from the explore release and 1,469 features from the expansion release. In addition to proteomic features, 55 clinical data fields were extracted to provide a comprehensive overview of each participant’s health and medical history.

### Data preprocessing

#### Defining cardiovascular disease

Our objective is to predict which participants will experience a primary cardiovascular disease (CVD) event within 10 years of recruitment. First, we excluded 4,119 participants with pre-existing CVD. Pre-existing CVD events were defined as coronary artery disease, ischemic stroke, and myocardial infarction using ICD 9/10 diagnostic codes from secondary care hospital episode statistics (**Table S3**), along with self-reported CVD events such as heart attack, angina and stroke (validated by clinicians). This filtering step resulted in a dataset with 50,057 participants.

Subsequently we defined our outcome of interest using a combination of hospital inpatient data, death register records (ONS), and self-reported medical conditions. We defined CVD as any of the following diagnoses: coronary artery disease, ischemic stroke, and myocardial infarction (**Table S3**), with study end-date of 1st February 2021. Participants not enrolled for the entire study duration (e.g. end-date < 10 years or participants that died of complications unrelated to CVD) were excluded from our analyses, which yielded a final set of 46,009 participants. Out of these, 3,287 were considered cases, 60% of which were men (**Table S4**).

#### Train and test data split

We divided all 46,009 participants into 10 groups. We ensured that the groups were roughly equally sized and matched by sex, age, ethnicity, assessment center and time until CVD diagnosis (**Supplementary** Figure 12). Specifically, we sorted male and female participants by date of diagnosis (if the participant is a case), or by date of recruitment (if the participant is a control). Participants were then iteratively assigned to each of the 10 groups. Groups were used for imputation, feature selection, and model training: we iteratively conducted each procedure on 9 of the 10 groups (“train set”), and only used the remaining group to measure performance (“test set”). This rendered 10 different splits of the data by taking different train and test sets, which allowed us to estimate the variance of our models’ performance across slightly different datasets.

#### Preprocessing of plasma proteomics measurements

We used Normalized Protein eXpression (NPX) values from the Olink platform, which employs Proximity Extension Assay technology (Sun, et al., 2023). This technology uses pairs of antibodies attached to unique oligonucleotides, binding specifically to target proteins and allowing for precise and sensitive plasma protein quantification. NPX values, presented on a log-2 scale, served as the quantification unit. Five participants with invalid data entries were excluded, particularly those lacking available normalized protein expression data.

#### Encoding clinical fields

We selected 55 clinical fields that were potentially useful predictors of CVD complications (**Table S5**). We excluded fields: (1) describing complications of CVD; (2) recorded after the initial assessment; (3) with identical values across all samples; or (4) with values missing in more than 99% of the participants. Subsequently, we encoded the selected fields according to their value type.^1^ Continuous fields were left unchanged. Categorical fields with single values, where a single answer is selected from a coded list, were also used as they were, except for two fields with relatively large sets of values, specifically, the assessment center (field 54) and Ethnic background (field 21000), which were mapped to smaller value sets. Fields with multiple values, where sets of answers are selected from a coded list, such as Qualifications (field 6138) and Illnesses of father (field 20107), underwent a one-hot encoding, resulting in one binary feature for each possible value. After encoding, the 55 fields were transformed into 173 features.

#### Imputation of clinical features

The 173 clinical features exhibited various degrees of missingness (**Table S5**): some had no missing values, like genetic sex, age at assessment date, or at recruitment; while others have missingness rates ranging from 0.18% (e.g., smoking status, alcohol intake frequency) to over 90% (e.g., number of cigarettes). Missingness rates were correlated within feature groups. For example, missingness for white blood cell count is identical to missingness for red blood cell count and hemoglobin concentration, likely indicating that the test used to assess these measures was not performed or invalid.

Crucially, none of the clinical features have values missing completely at random (Rubin, 1976). For instance, whether a value is missing for a given participant in the private healthcare feature (missingness rate of 65.65%) is highly dependent on values of features such as assessment center and deprivation index for that participant. In such cases, imputation with the mean or the median is not advised. Even state-of-the-art imputation methods such as MissForest (Stekhoven & Buhlmann, 2012) may cause unexpected problems (Chen, Tan, Chajewska, Rudin, & Caruna, 2023). Hence, we only imputed features with a low missingness rate (<1.5%), reducing the potential impact of incorrect imputations. For features which had a missingness rate above 1.5%, we replaced missing entries with a special value indicating “unknown.”

For features with a missingness rate below 1.5%, a Random Forest Regressor was utilized to model each feature with missing values as a function of other clinical features that exhibited low missingness. The imputation process was carried out through multiple iterative rounds, adhering to an iterated round-robin approach. Specifically, at each step within an iteration, one feature column with missing values was designated as the output variable, while the remaining feature columns were treated as input variables. It is worth noting that the imputer was trained exclusively on the training dataset to prevent data leakage and ensure the generalizability of the imputed values. This procedure was systematically applied to each feature requiring imputation. The iterative nature of the process allowed for refinement of the imputed values, with the algorithm proceeding for a maximum of 10 rounds or until a predefined stopping criterion was met. The stopping criterion in our study was set to trigger when the difference in imputed values between consecutive rounds became negligible, ensuring convergence to reliable estimates. For each split, the imputer, once trained on the training dataset, was then used to fill in missing values in both the training and the test datasets of that split. This ensured that the same imputation model was used across both datasets within each split, maintaining consistency in data handling and preserving the integrity of the imputation process.

### EBM models

In this manuscript, we focus on Explainable Boosting Machines (EBMs), an interpretable and highly performant model (Lou, Caruana, & Gehrke, 2013). In EBM models, the response variable depends linearly on unknown smooth functions of predictor variables (shape functions), with a link function, e.g., identity function for regression and the logistic function for classification. EBMs use bagged ensembles of boosted depth-restricted trees to represent the shape functions. We used the implementation in Python’s interpret package (Nori, Jenkins, Koch, & Caruana, 2019).

#### EBM feature selection

To minimize overfitting and utilize the most predictive proteins to train our final models, we performed a feature selection step. We trained 11 EBM models on the train set (Train and test data split) and all the proteins in the panel: one model on the outcome; three additional models that predict the phenotype at 3 different time horizons (3, 5, and 15 years); one model on a subset of samples in which cases are over-represented (all cases, only 20% of the controls); one model for each sex; one model for each age split (split at the median age of 58); and one model for each BMI split (split at the median BMI of 26.6).

We then calculated feature importance and selected the top 70 features with the highest feature importance for each model. The union of these features was then used to train the models. This yielded between 288 and 330 proteins across the 10 data splits (Train and test data split). The selected proteins were consistent across the splits: 248 were selected in at least half of the splits, and 138 appeared in all the splits.

We used machine learning to predict the outcome from all the clinical features and the selected proteins from the feature selection step.

#### EBM hyperparameter tuning

For hyperparameter tuning, we first split the training set into train (80%) and validation (20%) sets. We trained models with different hyperparameter configurations on the train portion of the training set and chose the optimal configuration based on the performance of these models on the validation set. The test set was not used in the optimization to avoid data leakage and overfitting. We used the Bayesian optimization algorithm (Snoek, Larochelle, & Adams, 2012) implemented in the hyperopt python package (Bergstra, Yamins, & Cox, 2013) to propose the series of tries leading to finding the optimal configuration. This algorithm allows us to find the optimal or near-optimal configuration in significantly fewer tries than exhaustive grid search and with higher probability than random search of the configuration space. We average hyperparameter settings from top M tries from each fold. We use M=10% of all tries.

#### EBM feature importance

In EBM models, feature importance values are calculated as mean absolute values of the logit function. To calculate the number, we look up the logit contribution for each feature for each training sample and take the absolute value of the logit, then we average those absolute values across all samples for each feature.

### State-of-the-art Models

#### Clinical scores

We compared our proteomics-only models for CVD risk prediction with three established clinical scores: PCE (Goff, et al., 2014), QRISK3 (Hippisley-Cox, Coupland, & Brindle, 2017) and SCORE2 (Graham, Di Angelantonio, Huculeci, & European Society of Cardiology’s Cardiovascular Ri, 2022). The Pooled Cohort Equations (PCE) provide sex- and race-specific estimations for 10-year atherosclerotic CVD risk, considering variables such as age, total cholesterol, high-density lipoprotein cholesterol, systolic blood pressure, diabetes mellitus, and current smoking status. QRISK3, a Cox proportional hazards model, predicts the 10-year risk of CVD in both men and women. The model considers 14 clinical factors, like age, ethnicity, and systolic blood pressure, along with eight additional risk factors, such as chronic kidney disease, and migraine. Last, the Systematic COronary Risk Evaluation (SCORE2) is a risk prediction model that estimates the 10-year CVD risk across different sexes, age groups, and regions, using factors like age, sex, smoking status, history of diabetes mellitus, systolic blood pressure, and total- and HDL-cholesterol.

#### Gradient boosting decision trees

We established a machine learning-based baseline, based on gradient boosting decision trees. Such models are a common choice for high-dimensional tabular data due to their speed, flexibility to handle non-linear relationships and high classification performance. Specifically, we used the LightGBM framework (Ke, et al., 2017). To extract interpretations from them, we used SHAP (SHapley Additive exPlanations) values, a game-theoretic approach used to explain the output of machine learning models. They provide insights into how each feature contributes to the prediction of a specific instance, allowing for a detailed understanding of the model’s decision-making process. SHAP values provide local interpretations, i.e., the contribution of each feature towards the individual prediction. To compute global interpretation, we took the absolute value of the feature contribution and averaged over all the training points in a split to get the global mean SHAP for each split.

We implemented a feature selection process to improve model performance and interpretability for LightGBM models. Initially, we trained our LightGBM models using the complete set of clinical and proteomic features based on the training data for each split. Subsequently, we computed the SHAP values to evaluate the global importance of each feature. The top 10% of features demonstrating the highest global importance, as determined by their SHAP values, were retained, and used to train the feature selected LightGBM model for each split. Notably, when testing the EBM-selected features with the LightGBM model, a slight decrease in predictive performance was observed compared to using the features selected through the abovementioned SHAP-value approach.

To search the hyperparameter space for the best performing model, we used the Azure Machine Learning’s AutoML framework. It uses a combination of Bayesian optimization and collaborative filteringto search for the optimal feature transformations and hyperparameter choices. For each of the 10 splits, we utilized the train portion of the data to tune hyperparameters. 4-fold cross validation was performed on the train portion and the best hyperparameter configuration was the one having highest mean dev AUROC. The process was repeated for each split leading to 10 separate best models for each split.

We also used SHAP values to understand the feature importance of the final models. There may be slight variations in feature importance and their relative ranking across splits. To ensure further robustness to global feature importance, we finally took the mean across splits and reported those top features.

### Model evaluation

We split the data into ten parts (Train and test data split), and use nine for data mining, and the remaining one for performance evaluation. We repeated this procedure ten times, obtaining ten unbiased measurements of performance. We evaluated the metrics above on the whole cohort, and across multiple stratifications, including sex, age and self-reported ethnicity.

We used two metrics of model performance: the area under the receiver operating characteristic curve (AUROC) and the area under the precision-recall curve (AUPRC). The AUROC measures the ability of a model to discriminate between positive and negative classes. A value of 1 indicates perfect prediction capability, while 0.5 indicates the model is not better than a random guess. The AUPRC measures the ability of the model to balance precision and recall, which is crucial in situations with highly imbalanced classes like the current setting. A perfect classifier has an AUPRC of 1, while the AUPRC of a random classifier would have a score around the proportion of positive classes.

Additionally, we used two additional metrics to assess the performance benefits of a new model: the net reclassification improvement (NRI) and integrated discrimination improvement (IDI). The NRI quantifies the net change in the number of individuals correctly reclassified as high-risk or low-risk when transitioning from Model 1 to Model 2. A positive NRI value indicates that Model 2 correctly reclassifies more individuals than Model 1, while a negative NRI value indicates that Model 2 incorrectly reclassifies more individuals than Model 1. The IDI, on the other hand, measures the overall improvement in the predictive model’s ability to distinguish between high-risk and low-risk individuals when transitioning from Model 1 to Model 2. A positive IDI value indicates that Model 2 is better at distinguishing between high-risk and low-risk individuals than Model 1, while a negative IDI value indicates that Model 2 is worse at distinguishing between high-risk and low-risk individuals than Model 1.

When evaluating 10-year disease risk, participants who developed CVD in the 11^th^ year after recruitment were more similar to participants who were positive within the first 15-years. Therefore, when computing the metrics, we decided to exclude the participants who developed CVD within 15 years.

### Quantifying feature importance differences by sex, age and BMI

To test if the observed differences in feature importance across sex, age, and BMI splits were due to limited sample size and stochastic noise, we generated a baseline experiment where we randomly split the population into two halves and trained a model using each half of the data.

Linear correlations between feature importance were calculated for the feature importance of the top 100 features. For non-overlapping features, we assigned a dummy feature importance as that of the 100^th^ feature in the group. To evaluate the linear correlation between the experimental group (data split by sex, age or BMI) against the random split, we performed the Fisher’s Z-transformation on both correlation coefficients and calculated the standard error for the differences in the two Z-scores. Then we calculated the P-value to assess if the correlation coefficient between the two splits deviates significantly from that of a random split. For instance, if there are significant differences in feature importance across sexes, we would anticipate that the splits between sexes would exhibit a significantly lower linear correlation of feature importance compared to that of the random split.

## Data Availability

No new data was generated in the present study

## Acknowledgements

This research has been conducted using the UK Biobank Resource under Application Numbers 53639 and 65851.

## Footnotes

1 https://biobank.ndph.ox.ac.uk/ukb/help.cgi?cd=value_type

## References

Adela, R., & Banerjee, S. K. (2015). GDF-15 as a Target and Biomarker for Diabetes and Cardiovascular Diseases: A Translational Prospective. J Diabetes Res, 2015, 490842.

Avram, R. (2023). Revolutionizing cardiovascular risk prediction in patients with chronic kidney disease: machine learning and large-scale proteomic risk prediction model lead the way. Eur Heart J, 44(23), 2111–2113.

Bergstra, J., Yamins, D., & Cox, D. (2013). Making a Science of Model Search: Hyperparameter Optimization in Hundreds of Dimensions for Vision Architectures. Proc. of the 30th International Conference on Machine Learning (ICML 2013).

Bycroft, C., Freeman, C., Petkova, D., Band, G., Elliott, L. T., Sharp, K.,…Marchini, J. (2018). The UK Biobank resource with deep phenotyping and genomic data. Nature, 562(7726), 203–209.

Cao, Z., Jia, Y., & Zhu, B. (2019). BNP and NT-proBNP as Diagnostic Biomarkers for Cardiac Dysfunction in Both Clinical and Forensic Medicine. Int J Mol Sci, 20(8), 1820.

Charo, F., & Taub, R. (2011). Anti-inflammatory therapeutics for the treatment of atherosclerosis. Nat Rev Drug Discov, 10(5), 365–376.

Chen, W., Schilperoort, M., Cao, Y., Shi, J., Tabas, I., & Tao, W. (2022). Macrophage-targeted nanomedicine for the diagnosis and treatment of atherosclerosis. Nat Rev Cardiol, 19(4), 228–249.

Chen, Z., Tan, S., Chajewska, U., Rudin, C., & Caruna, R. (2023). Missing Values and Imputation in Healthcare Data: Can Interpretable Machine Learning Help? Conference on Health, Inference and Learning (CHIL). arXiv, 2304.11749.

Gadd, D. A., Hillary, R. F., Kuncheve, Z., Mangelis, T., Cheg, Y., Dissanayake, M., & Sun, B. B. (2023). Blood protein levels predict leading incident diseases and mortality in UK Biobank. medRxiv, 2023-05.

Gow, D. C., Lloyd-Jones, D. M., Bennett, G., Coady, S., D’Agostino, R. B., Gibbons, R.,…American College of Cardiology/American Heart Asso, G. (2014). 2013 ACC/AHA guideline on the assessment of cardiovascular risk: a report of the American College of Cardiology/American Heart Association Task Force on Practice Guidelines. Circulation, 129(25 Suppl 2), S49–73.

Goncalves, I., Bengtsson, E., Colhoun, H. M., Shore, A. C., Palombo, C., Natali, A.,…Consortium, S. (2015). Elevated Plasma Levels of MMP-12 Are Associated With Atherosclerotic Burden and Symptomatic Cardiovascular Disease in Subjects With Type 2 Diabetes. Arterioscler Thromb Vasc Biol, 35(7), 1723–1731.

Graham, I. M., Di Angelantonio, E., Huculeci, R., & European Society of Cardiology’s Cardiovascular Ri, C. (2022). New Way to “SCORE” Risk: Updates on the ESC Scoring System and Incorporation into ESC Cardiovascular Prevention Guidelines. Curr Cardiol Rep, 24(11), 1679–1684.

Grinsztajn, L., Oyallon, E., & Varoquaux, G. (2022). Why do tree-based models still outperform deep learning on typical tabular data? Advances in Neural Information Processing Systems, 35, pp. 507–520.

Gudgeon, J., Marin-Rubio, J. L., & Trost, M. (2022). The role of macrophage scavenger receptor 1 (MSR1) in inflammatory disorders and cancer. Front Immunol, 13, 1012002.

Han, W. K., Bailly, V., Abichandani, R., Thadhani, R., & Bonventre, J. V. (2002). Kidney Injury Molecule-1 (KIM-1): a novel biomarker for human renal proximal tubule injury. Kidney Int, 62(1), 237–244.

Hastie, T., & Tibshirani, R. (1987). Generalized Additive Models: Some Applications. Journal of the American Statistical Association, 82(398).

Helgason, H., Eiriksdottir, T., Ulfarsson, M. O., Choudhary, A., Lund, S. H., Ivarsdottir, E. V.,…Stefansson, K. (2023). Evaluation of Large-Scale Proteomics for Prediction of Cardiovascular Events. JAMA, 330(8), 725–735.

Hippisley-Cox, J., Coupland, C., & Brindle, P. (2017). Development and validation of QRISK3 risk prediction algorithms to estimate future risk of cardiovascular disease: prospective cohort study. BMJ, 357, j2099.

Huang, B., Huang, W., Allen, J. C., Sun, L., Goh, H. J., Kong, S. C.,…Yeo, K. K. (2022). Prediction of subclinical atherosclerosis in low Framingham risk score individuals by using the metabolic syndrome criteria and insulin sensitivity index. Front Nutr, 9, 979208.

Ke, G., Meng, Q., Finley, T., Wang, T., Chen, W., Ma, W.,…Liu, T. Y. (2017). Lightgbm: A highly ewicient gradient boosting decision tree. Advances in neural information processing systems, 30, pp. 1–9.

Libby, P. (2021). The changing landscape of atherosclerosis. Nature, 592(7855), 524–533.

Lou, Y., Caruana, R., & Gehrke, J. (2013). Intelligible Models for Classification and Regression. Proceedings of the 18th ACM SIGKDD International Conference on Knowledge Discovery and Data Mining.

Lundberg, S. M., & Lee, S. I. (2017). A unified approach to interpreting model predictions. Advances in neural information processing systems, (pp. 4768–4777).

Ma, M., Hou, C., & Liu, J. (2023). Ewect of PCSK9 on atherosclerotic cardiovascular diseases and its mechanisms: Focus on immune regulation. Front Cardiovasc Med, 10, 1148486.

Newby, A. C. (2016). Metalloproteinase production from macrophages - a perfect storm leading to atherosclerotic plaque rupture and myocardial infarction. Exp Physiol, 101(11), 1327–1337.

Nightingale Health Biobank Collaborative Group, Barrett, J. C., Esko, T., Fischer, K., Jostins-Dean, L., Jousilahti, P., & Estonian Biobank Research Team. (2023). Metabolomic and genomic prediction of common diseases in 477,706 participants in three national biobanks. medRxiv, 2023-06.

Nori, H., Jenkins, S., Koch, P., & Caruana, R. (2019). InterpreML: A Unified Framework for Machine Learning Interpretability. arXiv, 1909.09223.

Nurmohamed, N. S., Kraaijenhof, J. M., Mayr, M., Nicholls, S. J., Koenig, W., Catapano, A. L., & Stroes, E. S. (2023). Proteomics and lipidomics in atherosclerotic cardiovascular disease risk prediction. Eur Heart J, 44(18), 1594–1607.

Rubin, D. (1976). Inference and Missing Data. Biometrika, 63(3), 581–592.

Sheng, W., Ji, G., & Zhang, L. (2022). Role of macrophage scavenger receptor MSR1 in the progression of non-alcoholic steatohepatitis. Front Immunol, 13, 1050984.

Singh, S. S., Pilkerton, C. S., Shrader, C. D., & Frisbee, S. J. (2018). Subclinical atherosclerosis, cardiovascular health, and disease risk: is there a case for the Cardiovascular Health Index in the primary prevention population? BMC Public Health, 18(1), 429.

Snoek, J., Larochelle, H., & Adams, R. P. (2012). Practical Bayesian Optimization of Machine Learning Algorithms. Advances in Neural Information Processing Systems, 25, pp. 1–9.

Stekhoven, D. J., & Buhlmann, P. (2012). MissForest--non-parametric missing value imputation for mixed-type data. Bioinformatics, 28(1), 112–118.

Sun, B. B., Chiou, J., Traylor, M., Benner, C., Hsu, Y., Richardson, T. G.,…Whelan, C. D. (2023). Plasma proteomic associations with genetics and health in the UK Biobank. Nature, 622(7982), 329–338.

Sun, B. B., Maranville, J. C., Peters, J. E., Stacey, D., Staley, J. R., Blackshaw, J.,…Butterworth, A. S. (2018). Genomic atlas of the human plasma proteome. Nature, 558(7708), 73–79.

Thompson, A., Gao, P., Orfei, L., Watson, S., Di Angelantonio, E., Kapto.,…Danesh, J. (2010). Lipoprotein-associated phospholipase A (2) and risk of coronary disease, stroke, and mortality: collaborative analysis of 32 prospective studies. Lancet, 375(9725), 1536–1544.

Traylor, M., Makela, K. M., Kilarski, L. L., Holliday, E. G., Devan, W. J., Nalls, M. A.,…Markus, H. S. (2014). A novel MMP12 locus is associated with large artery atherosclerotic stroke using a genome-wide age-at-onset informed approach. PLoS Genet, 10(7), e1004469.

Wang, D., Day, E. A., Townsend, L. K., Djordjevic, D., Jorgensen, S. B., & Steinberg, G. R. (2021). GDF15: emerging biology and therapeutic applications for obesity and cardiometabolic disease. Nat Rev Endocrinol, 592-607.

Williams, S. A., Ostrow, R., Hinterberg, M. A., Coresh, J., Ballantyne, C. M., Matsushita, K.,…Ganz, P. (2022). A proteomic surrogate for cardiovascular outcomes that is sensitive to multiple mechanisms of change in risk. Sci Transl Med, 14(639), eabj9625.

Wollert, K. C., Kempf, T., & Wallentin, L. (2017). Growth Diwerentiation Factor 15 as a Biomarker in Cardiovascular Disease. Clin Chem, 63(1), 140–151.

