## Supplementary figures and tables for "Interpretable Machine Learning Leverages Proteomics to Improve Cardiovascular Disease Risk Prediction and Biomarker Identification"

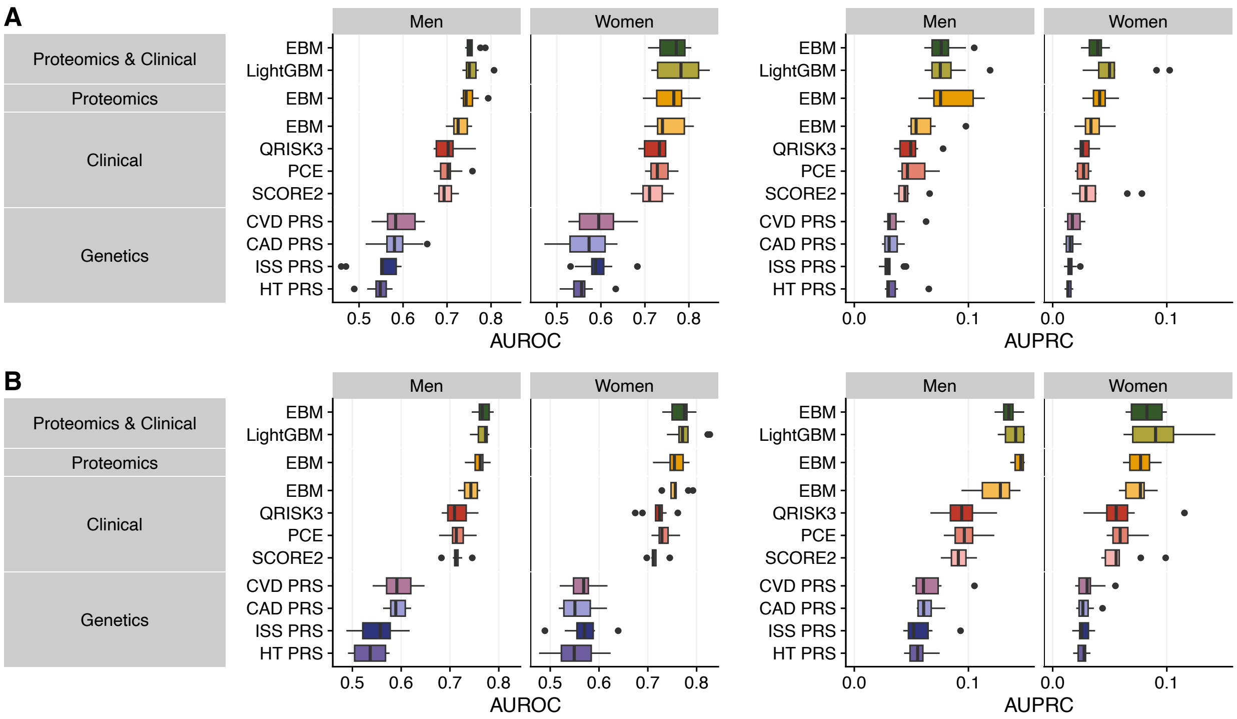


**Supplementary Figure 1**. Model performance for **(A)** 3-year and **(B)** 5-year atherosclerosis risk prediction using the AUROC (left) and the AUPRC (right).

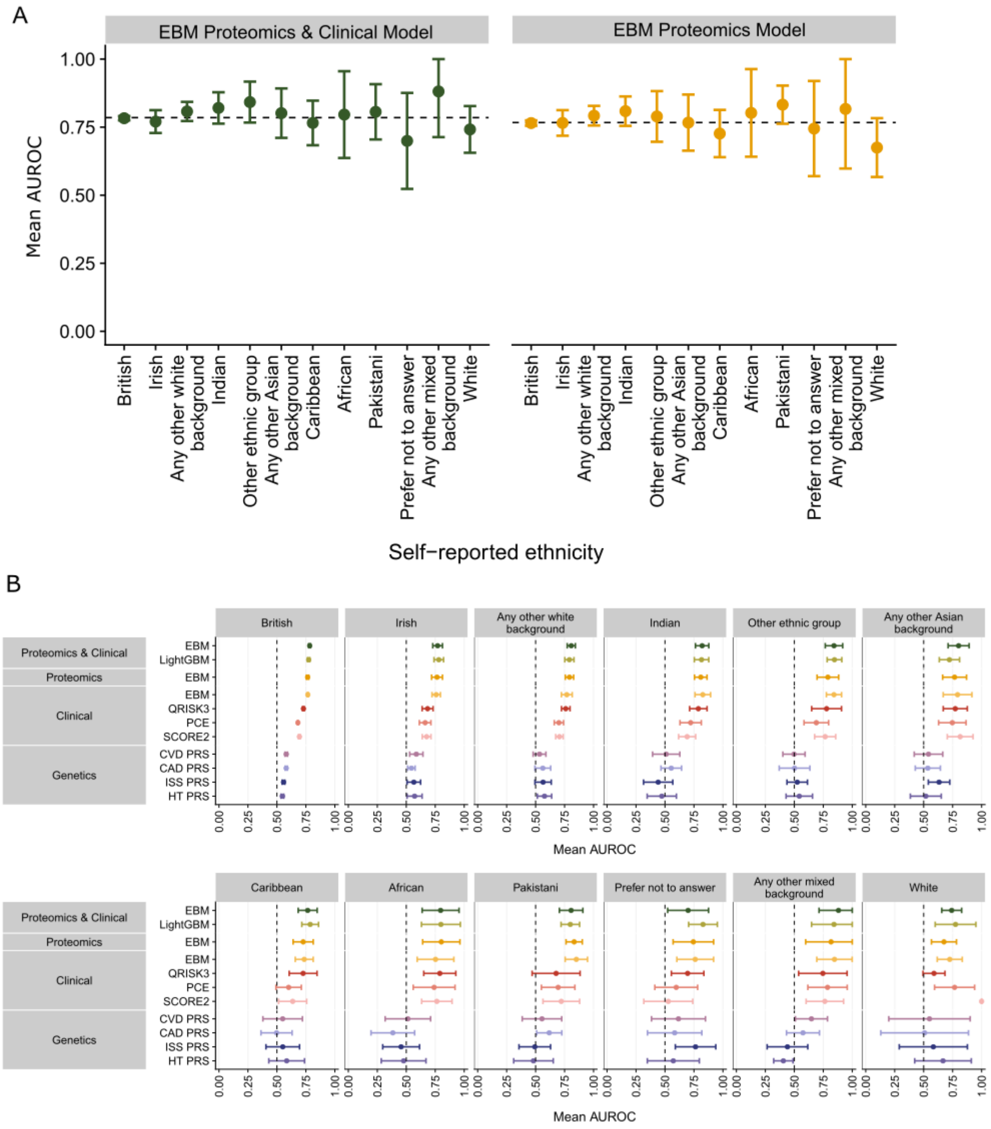


**Supplementary Figure 2. Consistent model performance across ethnicities. (A)** The performances of the EBM Proteomics & Clinical, and the EBM Proteomics models are consistent across self-reported ethnicities. **(B)** Overall, the EBM Proteomics & Clinical model outperforms other traditional models across ethnicities. Only ethnicities with at least five cases are reported. Error bars show the 95% confidence interval.

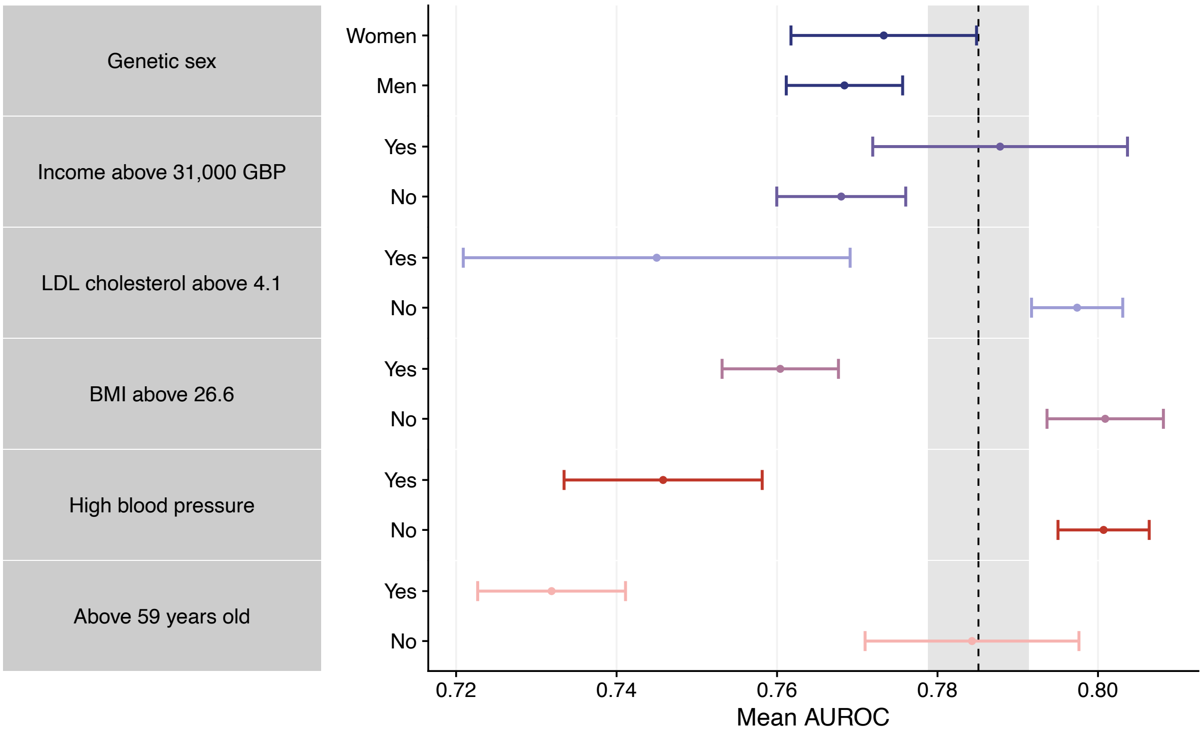


**Supplementary Figure 3. Model performance across various subpopulations.** The mean value is represented with a dot, and the 95% confidence interval with the error bars. The dashed line represents the mean AUROC across the entire cohort, and the gray shaded area shows its 95% confidence interval.

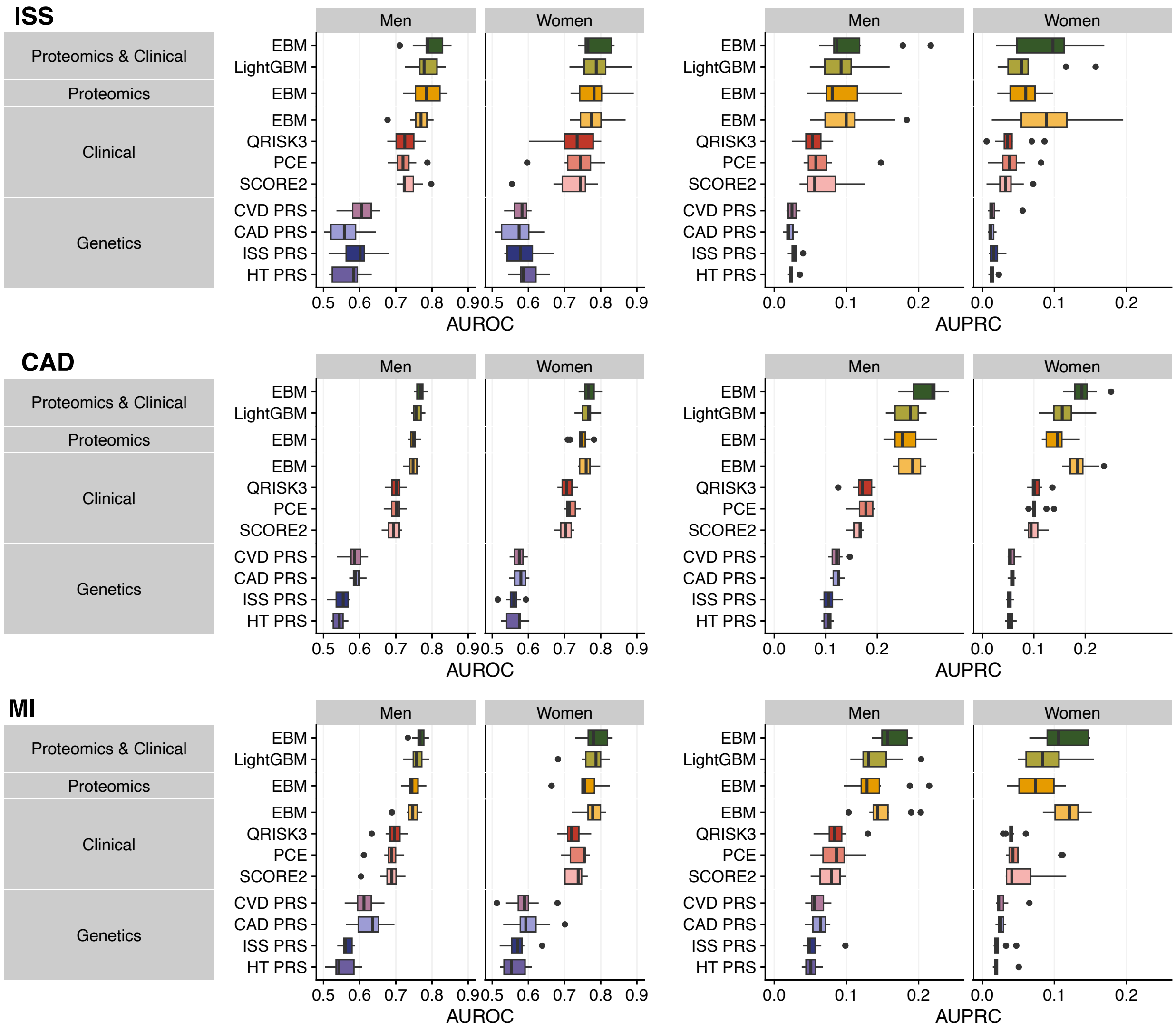


**Supplementary Figure 4.** Model performance in predicting three CVD outcomes within 10 years: ischemic stroke **(ISS)**, coronary artery disease **(CAD),** and myocardial infarction **(MI)**. The performance is represented by the AUROC on the left side and the AUPRC on the right side.

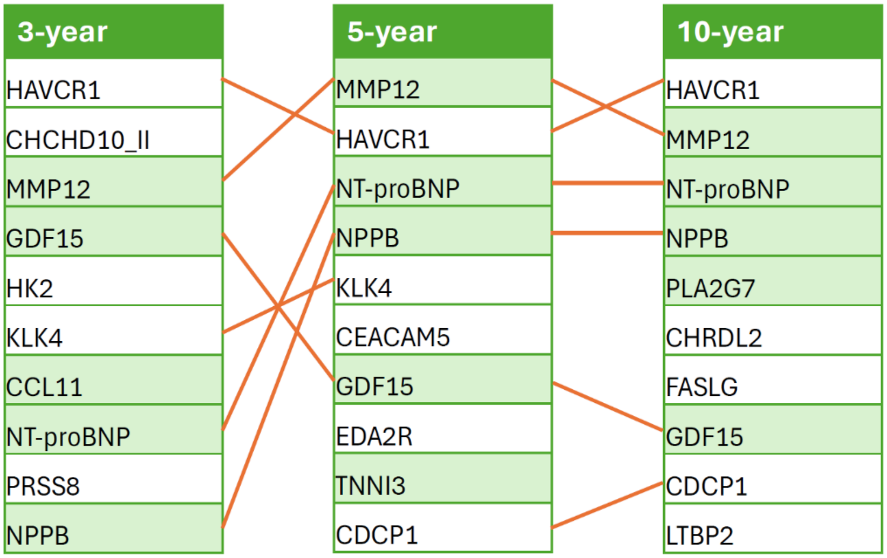


**Supplementary Figure 5**. Top 10 contributing proteins at different time horizons. The shared features are connected by orange lines. Notably, six of the top 10 features were shared between the 3-and 5-year predictions, as well as between the 5-year and 10-year predictions. Biomarkers that have been previously identified are highlighted with a green shade.

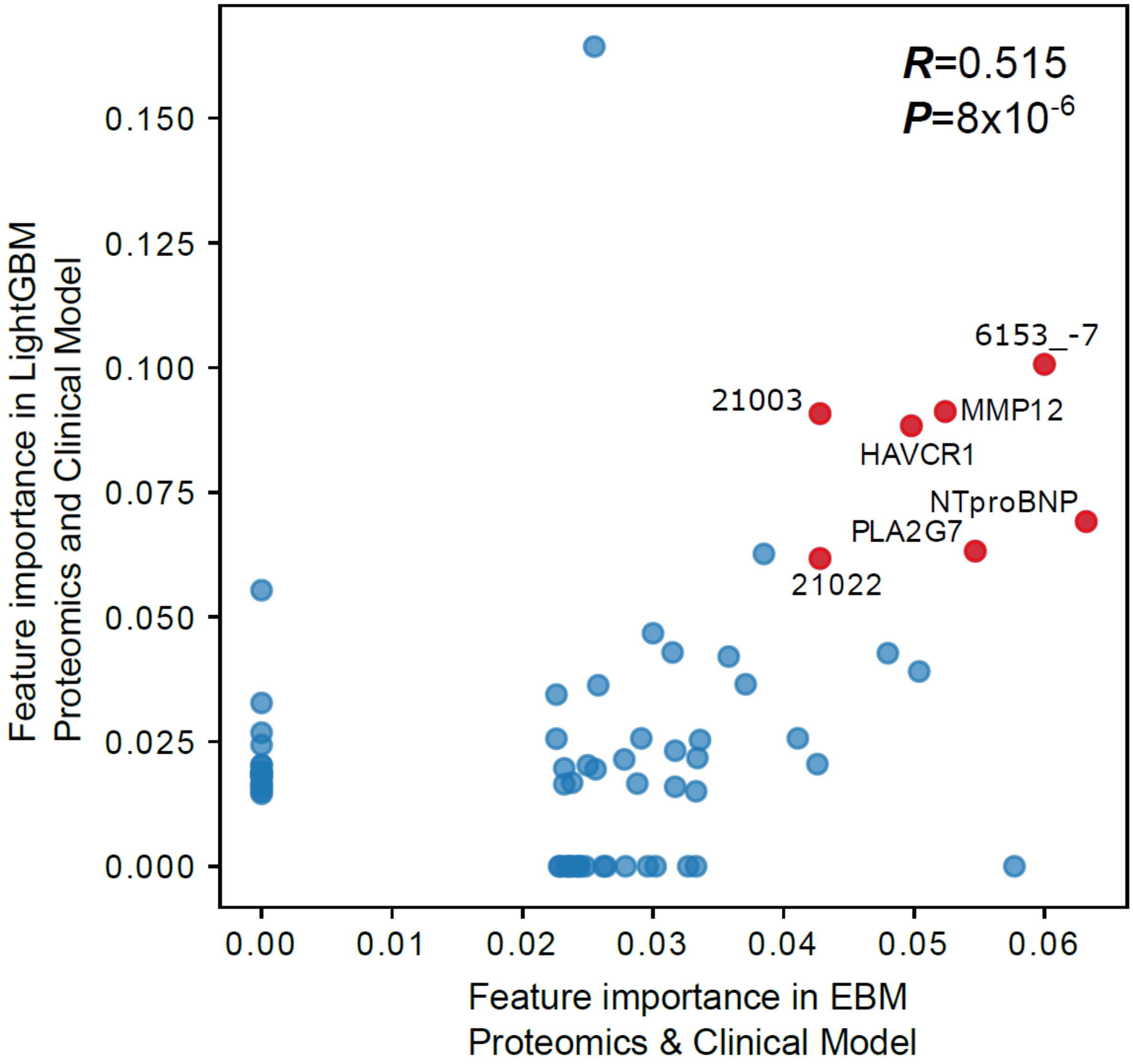


**Supplementary Figure 6**. SHAP values of the top 50 features in the LightGBM models. The seven features that overlap with the top 10 features in the EBM Proteomics & Clinical model are highlighted in red. 6153_-7: none of the above for “Medication for cholesterol, blood pressure, diabetes, or take exogenous hormones”. 21003: Age when attended assessment centre. 6150_-7: none of the above for “Vascular/heart problems diagnosed by doctor”. 21022: Age at recruitment.

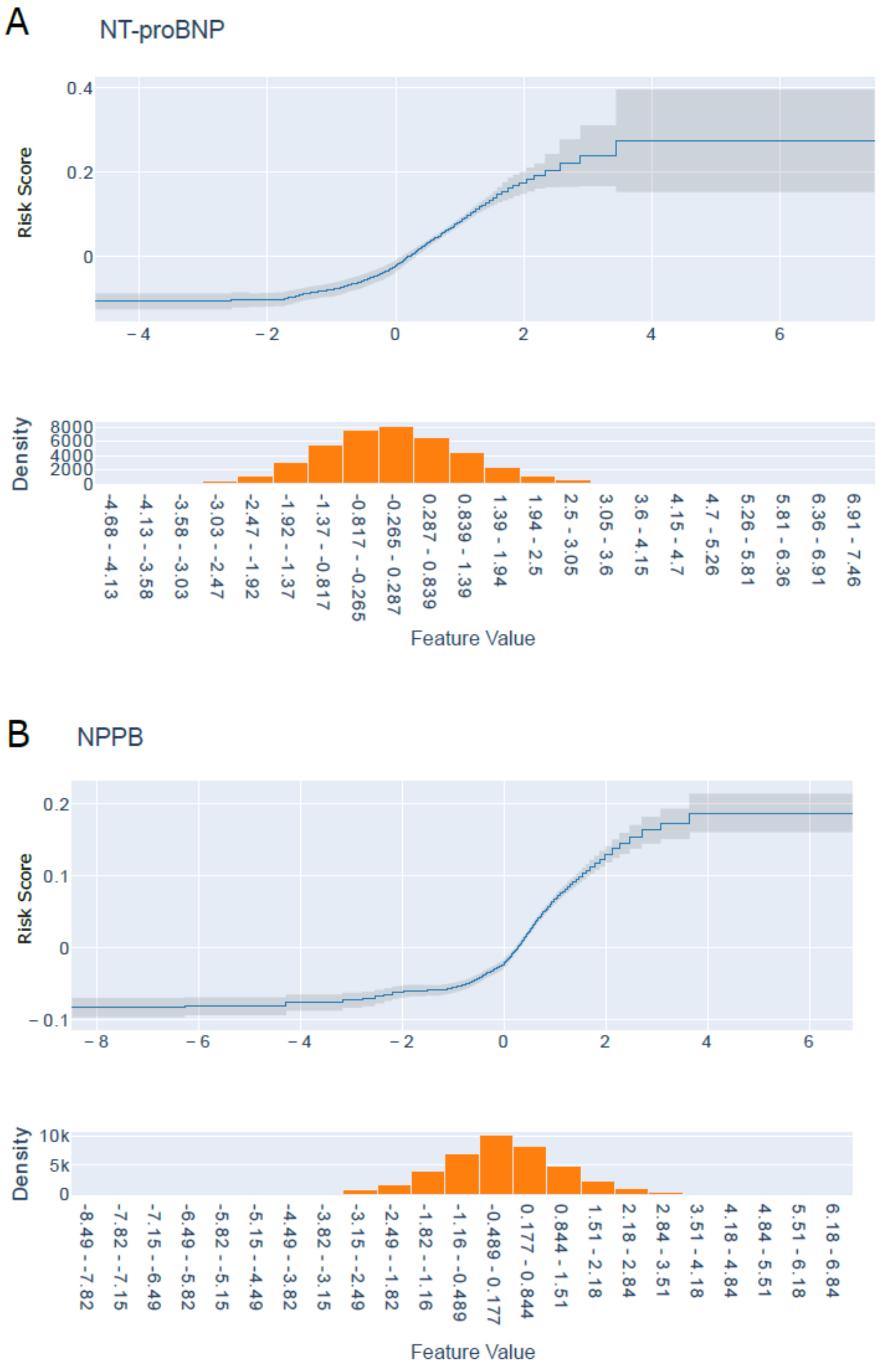


**Supplementary Figure 7. (A)** NT-proBNP and **(B)** NPPB, both involved in the natriuretic peptide system, show similar disease risk profile. Note the scale of the Y-axis on the panels were different, and NT-proBNP has a more profound impact than NPPB. The top panels of each subfigure display the risk score on the logarithmic scale for different feature values, with gray shaded regions representing the standard deviation for estimations. The bottom panels show histograms of the feature values across all participants.

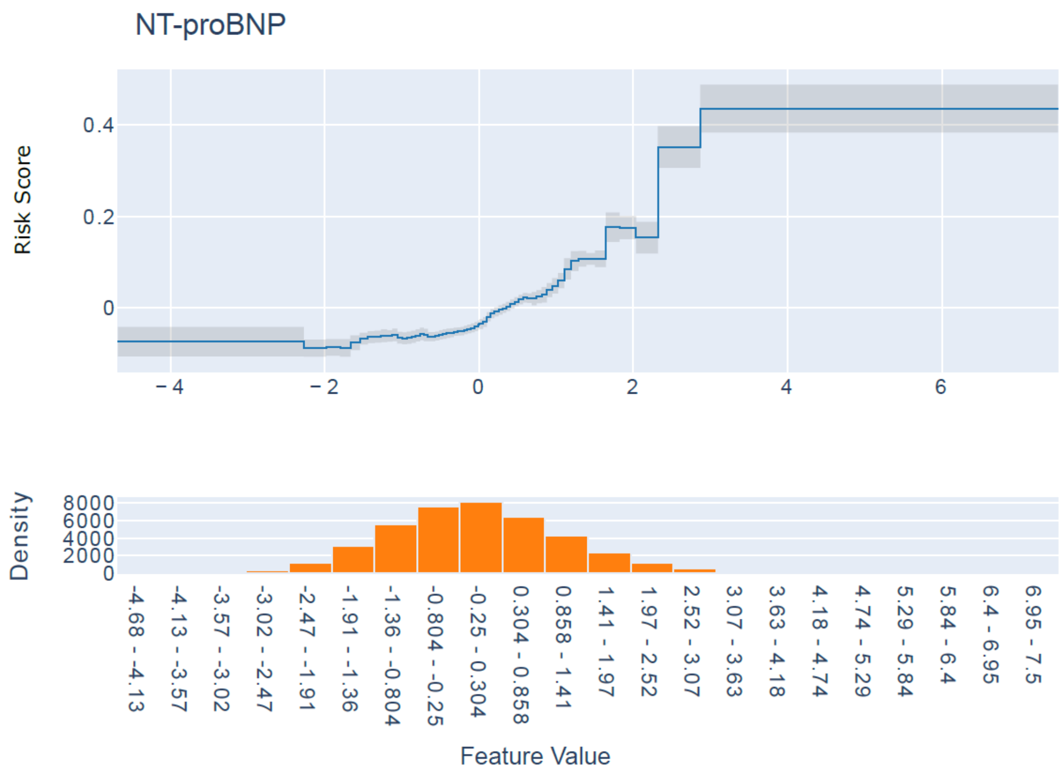


**Supplementary Figure 8.** Risk profile of NT-ProBNP in the EBM Proteomics & Clinical model. It shows a weaker impact compared to that of the EBM Proteomics model (Supplementary Figure 7A). The top panel displays the risk score on the logarithmic scale for different feature values, with gray shaded regions representing the standard deviation for estimations. The bottom panel shows histograms of the feature values across all participants.

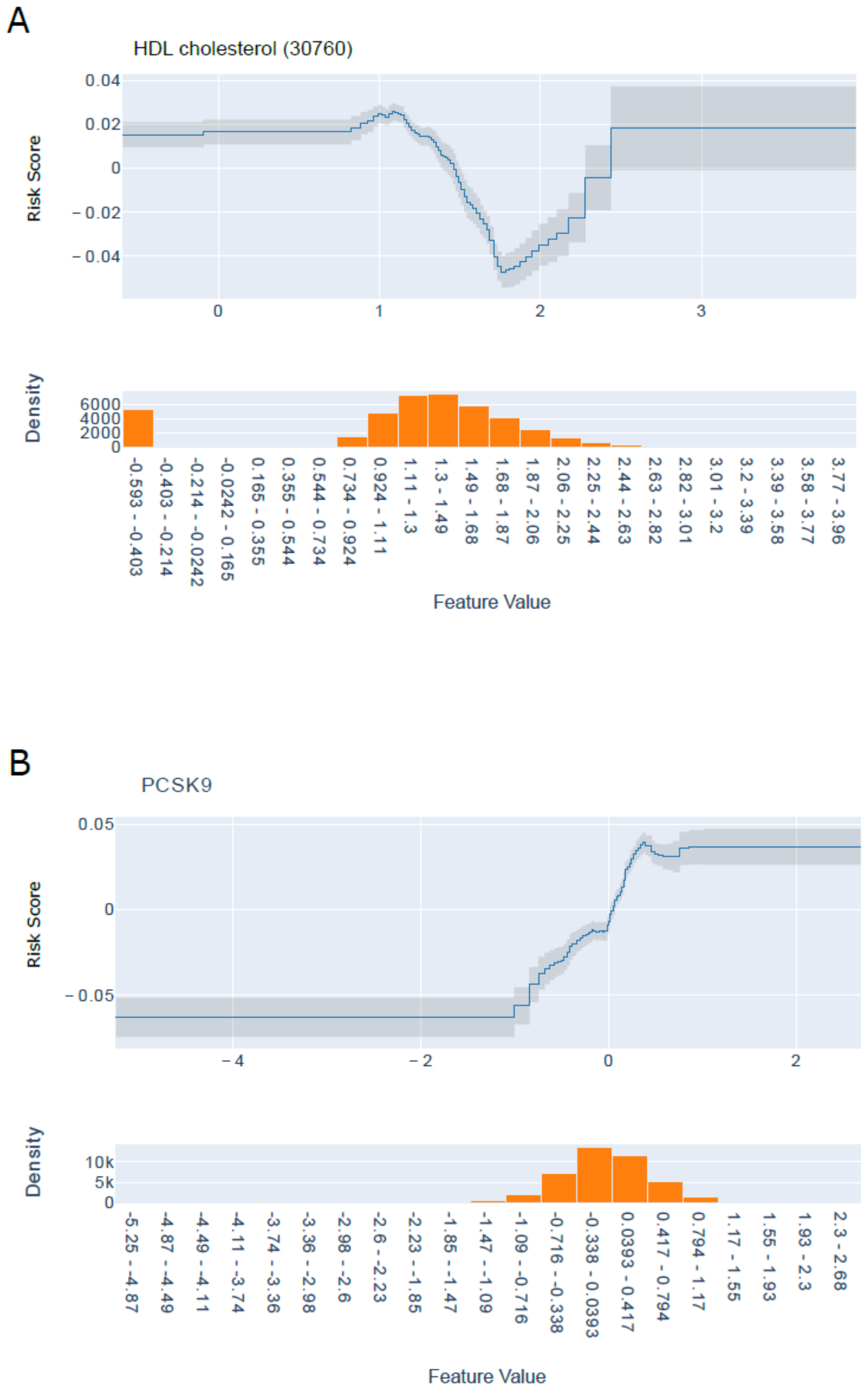


**Supplementary Figure 9.** Risk profiles of **(A)** HDL and **(B)** PCSK9 in the EBM Proteomics & Clinical model. The top panels of each subfigure display the risk score on the logarithmic scale for different feature values, with gray shaded regions representing the standard deviation for estimations. The bottom panels show histograms of the feature values across all participants.

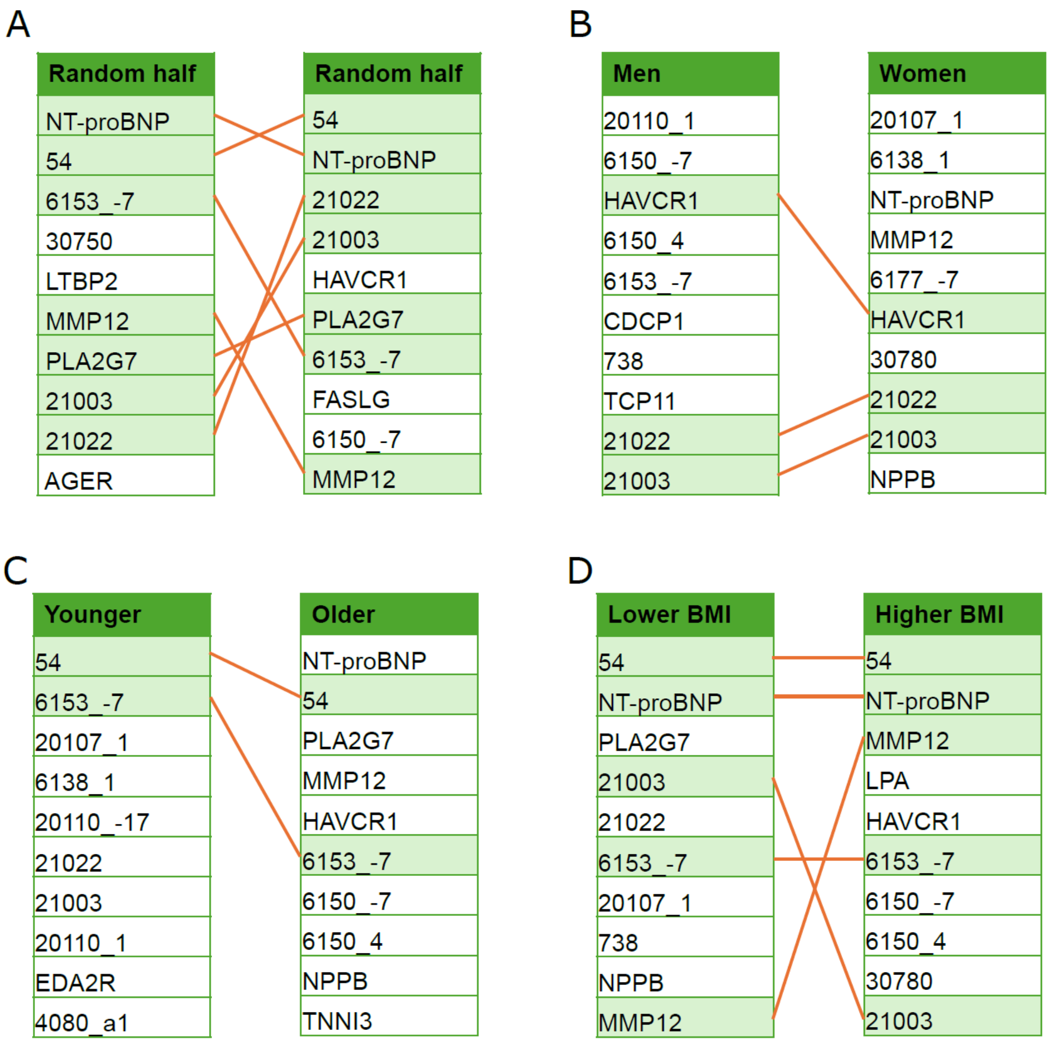


**Supplementary Figure 10.** The list of top 10 features is shown for **(A)** models built based on random split, **(B)** separate models built for different genders, **(C)** ages, and **(D)** BMIs, respectively, in the EBM Proteomics & Clinical model. We split age and BMI based on the median value. Shared features between splits were highlighted in green and linked by orange lines. Random splits share 7/10 features, while split by gender, age and BMI have 3, 2 and 5 shared top 10 features respectively. The feature encodings of the clinical features are listed in Table S3.

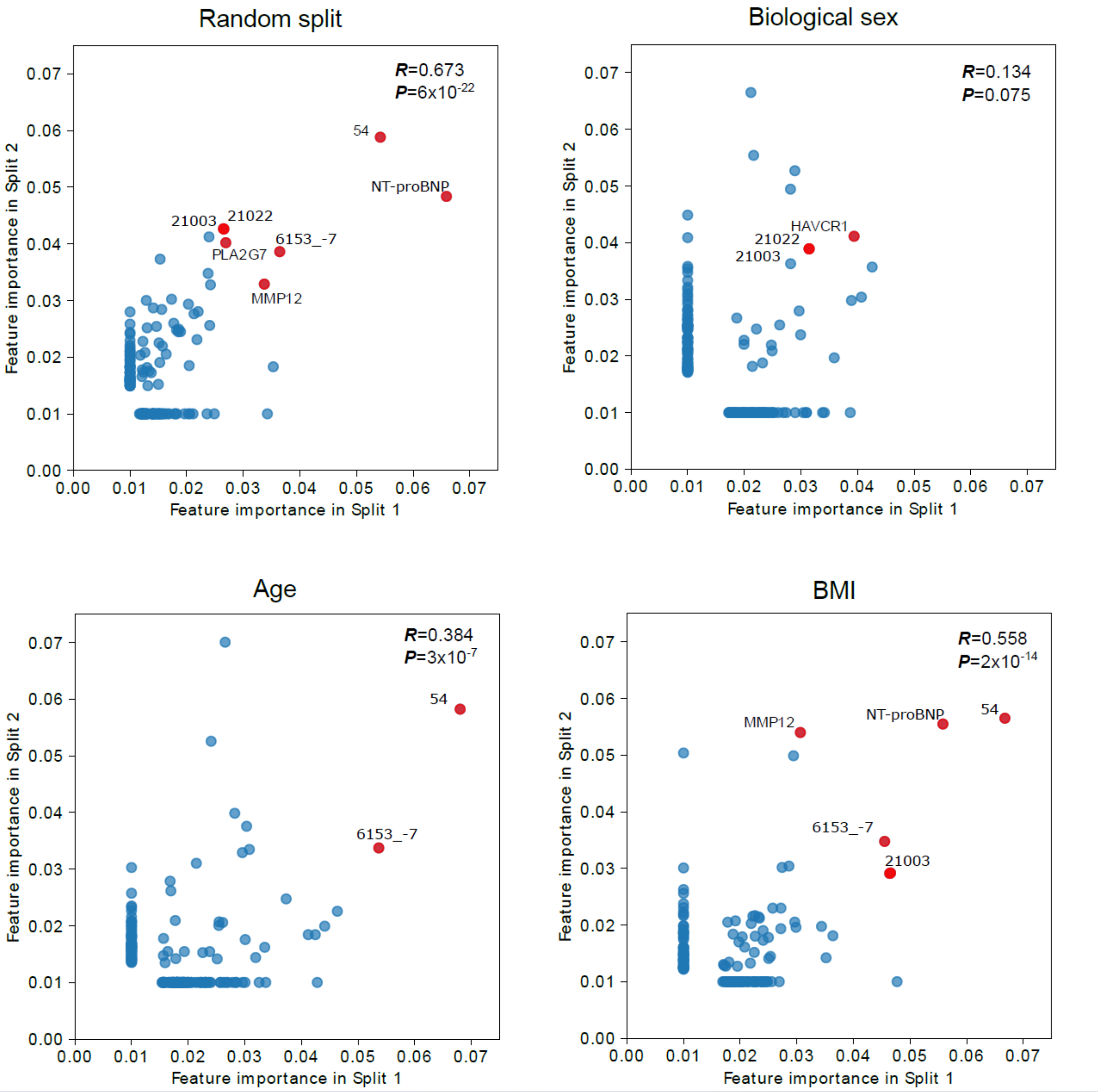


**Supplementary Figure 11.** The feature importance for Top 100 features were plotted against models based on the two data splits. Pearson's R and the associated P values are shown in the upper right corner. Shared top 10 features in both data splits are highlighted in red. Note that the clinical fields 21022 (Age at recruitment) and 21003 (Age when attended assessment center) were highly correlated, shown as overlapping dots with similar feature importance. The other displayed clinical fields were 54 (UK Biobank assessment center) and 6153_-7 (None of the above for "Medication for cholesterol, blood pressure, diabetes, or take exogenous hormones").

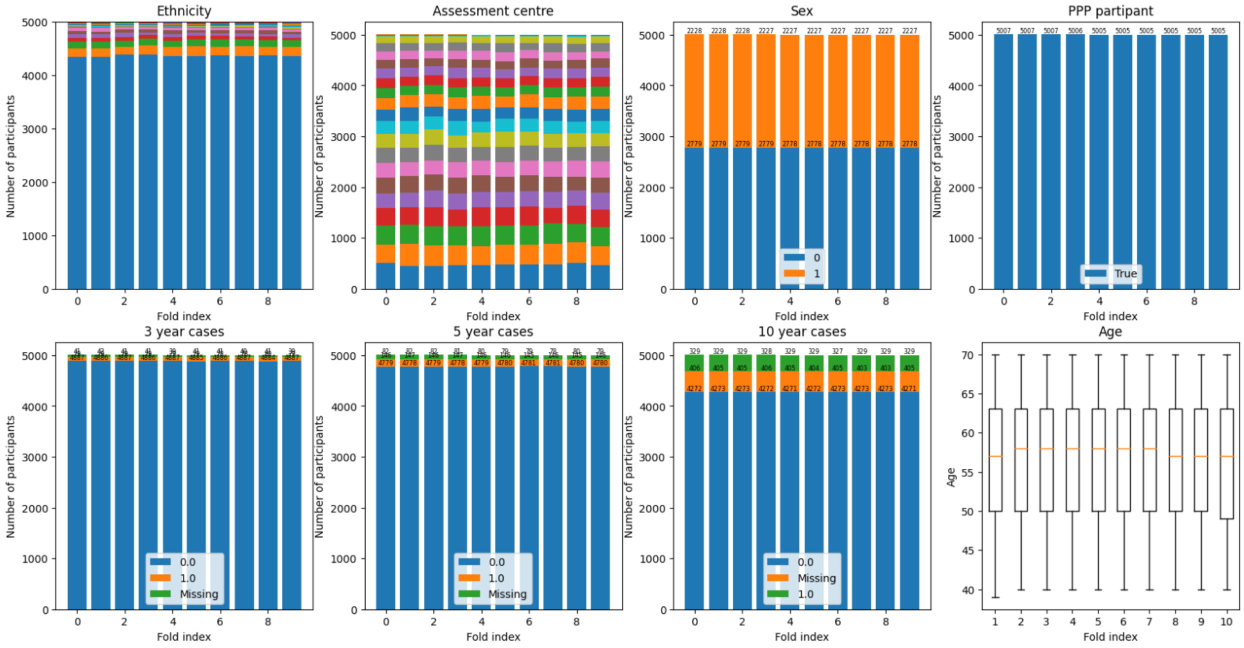


**Supplementary Figure 12**. Phenotypic characteristics of the participants in the different data splits were consistent.

**Table S1.** Comparison of discrimination and reclassification performance between various EBM models and QRISK3 over a 10-year time horizon.

| | | **A. QRISK3 vs. EBM Clinical** | **B. QRISK3 vs EBM Proteomics** | **C. QRISK3 vs EBM Proteomics & Clinical** | **D. EBM Proteomics vs EBM Proteomics & Clinical** |
|---|---|---|---|---|---|
| NRI | Index | 0.5908 | 0.5538 | 0.708 | 0.3657 |
| | 95% CI | (0.3566, 0.825) | (0.3157, 0.7918) | (0.4763, 0.9397) | (0.1445, 0.587) |
| | P-value | .0001 *** | .0001 *** | < .0001 **** | .0033 ** |
| IDI | Index | 0.0635 | 0.0555 | 0.0926 | 0.0329 |
| | 95% CI | (0.0213, 0.1057) | (0.0235, 0.0874) | (0.0474, 0.1379) | (0.0044, 0.0615) |
| | P-value | .0134 * | .0047 ** | .0004 *** | .0463 * |

P-value: *: P<.05; **: P<.01; ***: P<.001; ****: P<.0001

**Table S2.** Clinical features among the top 20 most important features in the EBM Proteomics & Clinical model. Note that no directionality is implied regarding how the predictive value influences CVD risk.

| Rank | Variable (field number) | Predictive value(s) (when multiple values are allowed) |
|---|---|---|
| 2 | Medication for cholesterol, blood pressure, diabetes, or take exogenous hormones (6153) | None of the above |
| 3 | Assessment center (54) | - |
| 6 | Illnesses of father (20107) | Heart disease |
| 8 | Illnesses of mother (20110) | Heart disease |
| 9 | Age when attended assessment center (21003) | - |
| 10 | Age at recruitment (21022) | - |
| 11 | Qualifications (6138) | College or University degree |
| 13, 14 | Vascular/heart problems diagnosed by doctor (6150) | High blood pressure, None of the above |
| 15 | Systolic blood pressure, automated reading (4080) | - |
| 16 | Glycated hemoglobin (HbA1c) (30750) | - |
| 17 | Medication for cholesterol, blood pressure or diabetes (6177) | Missing value |
| 18 | Sex (31) | - |
| 19 | LDL levels (30780) | - |

**Table S3.** Disease codes used to define atherosclerosis events.

| Phenotype | ICD-9 | ICD-10 | OPCS-4 | Self-reported |
|---|---|---|---|---|
| **Coronary artery disease** | 410, 411, 412 | I20, I21, I22, I23, I24, I25 | K40, K41, K42, K43, K44, K45, K46, K49, K501, K759 | 1075 |
| **Ischemic stroke** | 434, 436 | I63, I64 | - | 1583 |
| **Myocardial infarction** | 410, 411, 412 | I21, I22, I23, I24, I252 | - | 1075 |

**Table S4.** Count of cases at different time horizons for the entire cohort, and separately for men and women. The total number of participants is shown in parenthesis within the studied cohort.

| Time horizon | Total | Men | Women |
|---|---:|---:|---:|
| 3-year outcome | 789 (49,650) | 506 (22,035) | 283 (27,615) |
| 5-year outcome | 1,460 (49,254) | 922 (21,824) | 538 (27,430) |
| 10-year outcome | 3,287 (46,009) | 1,999 (20,269) | 1,288 (25,740) |

**Table S5.** Different clinical features exhibit various degrees of data missingness.

(Shared separately)

**Table S6.** Mean performance and 95% confidence interval of the models shown in Supplementary Figure 4. The best performing model in each column for each outcome is highlighted in bold.

**Table S6.1.** Area under the receiver operating characteristic curve

| Clinical outcome | Data | Model | Overall | Women | Men |
|---|---|---|---|---|---|
| Coronary artery disease | Proteomics & Clinical | EBM | **0.7846 ± 0.0055** | **0.7699 ± 0.0127** | **0.7673 ± 0.0078** |
| Coronary artery disease | Proteomics & Clinical | LightGBM | 0.7761 ± 0.0090 | 0.7612 ± 0.0133 | 0.7587 ± 0.0102 |
| Coronary artery disease | Proteomics | EBM | 0.7660 ± 0.0078 | 0.7468 ± 0.0137 | 0.7498 ± 0.0073 |
| Coronary artery disease | Clinical | EBM | 0.7703 ± 0.0041 | 0.7595 ± 0.0123 | 0.7457 ± 0.0100 |
| Coronary artery disease | Clinical | QRISK3 | 0.7244 ± 0.0102 | 0.7074 ± 0.0114 | 0.6987 ± 0.0120 |
| Coronary artery disease | Clinical | PCE | 0.6935 ± 0.0080 | 0.7180 ± 0.0096 | 0.6997 ± 0.0112 |
| Coronary artery disease | Clinical | SCORE2 | 0.6919 ± 0.0078 | 0.7025 ± 0.0114 | 0.6935 ± 0.0118 |
| Coronary artery disease | Genetics | CVD PRS | 0.5782 ± 0.0116 | 0.5732 ± 0.0102 | 0.5860 ± 0.0151 |
| Coronary artery disease | Genetics | CAD PRS | 0.5829 ± 0.0074 | 0.5774 ± 0.0118 | 0.5907 ± 0.0082 |
| Coronary artery disease | Genetics | ISS PRS | 0.5497 ± 0.0116 | 0.5580 ± 0.0131 | 0.5482 ± 0.0143 |
| Coronary artery disease | Genetics | HT PRS | 0.5459 ± 0.0090 | 0.5552 ± 0.0223 | 0.5418 ± 0.0104 |
| Ischemic stroke | Proteomics & Clinical | EBM | **0.8020 ± 0.0200** | **0.7981 ± 0.0325** | **0.7937 ± 0.0269** |
| Ischemic stroke | Proteomics & Clinical | LightGBM | 0.7922 ± 0.0178 | 0.7831 ± 0.0325 | 0.7877 ± 0.0249 |
| Ischemic stroke | Proteomics | EBM | 0.7890 ± 0.0176 | 0.7807 ± 0.0312 | 0.7838 ± 0.0263 |
| Ischemic stroke | Clinical | EBM | 0.7778 ± 0.0169 | 0.7758 ± 0.0284 | 0.7640 ± 0.0227 |
| Ischemic stroke | Clinical | QRISK3 | 0.7255 ± 0.0155 | 0.7310 ± 0.0384 | 0.7257 ± 0.0206 |
| Ischemic stroke | Clinical | PCE | 0.6859 ± 0.0171 | 0.7341 ± 0.0368 | 0.7225 ± 0.0208 |
| Ischemic stroke | Clinical | SCORE2 | 0.7298 ± 0.0165 | 0.7193 ± 0.0427 | 0.7360 ± 0.0186 |
| Ischemic stroke | Genetics | CVD PRS | 0.5629 ± 0.0306 | 0.5445 ± 0.0445 | 0.5788 ± 0.0380 |
| Ischemic stroke | Genetics | CAD PRS | 0.5294 ± 0.0294 | 0.5329 ± 0.0459 | 0.5290 ± 0.0414 |
| Ischemic stroke | Genetics | ISS PRS | 0.5883 ± 0.0192 | 0.5845 ± 0.0296 | 0.5931 ± 0.0286 |
| Ischemic stroke | Genetics | HT PRS | 0.5655 ± 0.0231 | 0.5734 ± 0.0400 | 0.5608 ± 0.0308 |
| Myocardial infarction | Proteomics & Clinical | EBM | **0.7937 ± 0.0108** | **0.7874 ± 0.0214** | **0.7667 ± 0.0114** |
| Myocardial infarction | Proteomics & Clinical | LightGBM | 0.7834 ± 0.0147 | 0.7768 ± 0.0267 | 0.7551 ± 0.0153 |
| Myocardial infarction | Proteomics | EBM | 0.7739 ± 0.0141 | 0.7605 ± 0.0263 | 0.7486 ± 0.0120 |
| Myocardial infarction | Clinical | EBM | 0.7792 ± 0.0098 | 0.7779 ± 0.0174 | 0.7442 ± 0.0147 |
| Myocardial infarction | Clinical | QRISK3 | 0.7358 ± 0.0096 | 0.7232 ± 0.0180 | 0.6942 ± 0.0172 |
| Myocardial infarction | Clinical | PCE | 0.7071 ± 0.0096 | 0.7405 ± 0.0172 | 0.6837 ± 0.0184 |
| Myocardial infarction | Clinical | SCORE2 | 0.6934 ± 0.0172 | 0.7295 ± 0.0163 | 0.6831 ± 0.0212 |
| Myocardial infarction | Genetics | CVD PRS | 0.6008 ± 0.0186 | 0.5882 ± 0.0284 | 0.614 ± 0.0202 |
| Myocardial infarction | Genetics | CAD PRS | 0.6155 ± 0.0225 | 0.6042 ± 0.0302 | 0.6273 ± 0.0263 |
| Myocardial infarction | Genetics | ISS PRS | 0.5577 ± 0.0098 | 0.5531 ± 0.0298 | 0.5648 ± 0.0100 |
| Myocardial infarction | Genetics | HT PRS | 0.5525 ± 0.0165 | 0.5521 ± 0.0265 | 0.5558 ± 0.0216 |

**Table S6.2.** Area under the precision-recall curve

| Clinical outcome | Data | Model | Overall | Women | Men |
|---|---|---|---|---|---|
| Coronary artery disease | Proteomics & Clinical | EBM | **0.2591 ± 0.0125** | **0.1943 ± 0.0172** | **0.296 ± 0.0194** |
| Coronary artery disease | Proteomics & Clinical | LightGBM | 0.2176 ± 0.0125 | 0.1613 ± 0.0145 | 0.2519 ± 0.0186 |
| Coronary artery disease | Proteomics | EBM | 0.2157 ± 0.0165 | 0.1454 ± 0.0155 | 0.2581 ± 0.0214 |
| Coronary artery disease | Clinical | EBM | 0.2364 ± 0.0078 | 0.1872 ± 0.0167 | 0.2633 ± 0.0153 |
| Coronary artery disease | Clinical | QRISK3 | 0.1463 ± 0.0090 | 0.1048 ± 0.0086 | 0.171 ± 0.0135 |
| Coronary artery disease | Clinical | PCE | 0.1413 ± 0.0074 | 0.1052 ± 0.0092 | 0.1754 ± 0.0112 |
| Coronary artery disease | Clinical | SCORE2 | 0.1256 ± 0.0069 | 0.0987 ± 0.0092 | 0.162 ± 0.0071 |
| Coronary artery disease | Genetics | CVD PRS | 0.0833 ± 0.0033 | 0.0575 ± 0.0051 | 0.1213 ± 0.0078 |
| Coronary artery disease | Genetics | CAD PRS | 0.0845 ± 0.0031 | 0.0583 ± 0.0029 | 0.1218 ± 0.0059 |
| Coronary artery disease | Genetics | ISS PRS | 0.0747 ± 0.0039 | 0.0528 ± 0.0033 | 0.1064 ± 0.0080 |
| Coronary artery disease | Genetics | HT PRS | 0.0746 ± 0.0027 | 0.0541 ± 0.0041 | 0.1041 ± 0.0053 |
| Ischemic stroke | Proteomics & Clinical | EBM | **0.0991 ± 0.0202** | **0.0868 ± 0.0298** | **0.1105 ± 0.0308** |
| Ischemic stroke | Proteomics & Clinical | LightGBM | 0.0735 ± 0.0141 | 0.0587 ± 0.0176 | 0.0926 ± 0.0214 |
| Ischemic stroke | Proteomics | EBM | 0.0708 ± 0.0123 | 0.0578 ± 0.0153 | 0.0927 ± 0.0243 |
| Ischemic stroke | Clinical | EBM | 0.0948 ± 0.0245 | 0.0913 ± 0.0325 | 0.1029 ± 0.0272 |
| Ischemic stroke | Clinical | QRISK3 | 0.0416 ± 0.0073 | 0.0392 ± 0.0143 | 0.0529 ± 0.0106 |
| Ischemic stroke | Clinical | PCE | 0.0472 ± 0.0143 | 0.039 ± 0.0131 | 0.0664 ± 0.0194 |
| Ischemic stroke | Clinical | SCORE2 | 0.052 ± 0.0110 | 0.0346 ± 0.0116 | 0.0677 ± 0.0184 |
| Ischemic stroke | Genetics | CVD PRS | 0.0183 ± 0.0031 | 0.0179 ± 0.0088 | 0.0251 ± 0.0043 |
| Ischemic stroke | Genetics | CAD PRS | 0.0158 ± 0.0024 | 0.0127 ± 0.0025 | 0.022 ± 0.0041 |
| Ischemic stroke | Genetics | ISS PRS | 0.0205 ± 0.0027 | 0.0175 ± 0.0047 | 0.0279 ± 0.0035 |
| Ischemic stroke | Genetics | HT PRS | 0.0176 ± 0.0014 | 0.014 ± 0.0025 | 0.0241 ± 0.0029 |
| Myocardial infarction | Proteomics & Clinical | EBM | **0.1472 ± 0.0106** | **0.113 ± 0.0202** | **0.1643 ± 0.0129** |
| Myocardial infarction | Proteomics & Clinical | LightGBM | 0.1117 ± 0.0104 | 0.0899 ± 0.0167 | 0.1297 ± 0.0159 |
| Myocardial infarction | Proteomics | EBM | 0.112 ± 0.0139 | 0.0749 ± 0.0176 | 0.1405 ± 0.0220 |
| Myocardial infarction | Clinical | EBM | 0.1385 ± 0.0131 | 0.1184 ± 0.0143 | 0.1503 ± 0.0178 |
| Myocardial infarction | Clinical | QRISK3 | 0.0705 ± 0.0088 | 0.0406 ± 0.0051 | 0.0845 ± 0.0135 |
| Myocardial infarction | Clinical | PCE | 0.0688 ± 0.0102 | 0.0547 ± 0.0186 | 0.0866 ± 0.0159 |
| Myocardial infarction | Clinical | SCORE2 | 0.0608 ± 0.0108 | 0.0543 ± 0.0176 | 0.0776 ± 0.0104 |
| Myocardial infarction | Genetics | CVD PRS | 0.0393 ± 0.0037 | 0.0287 ± 0.0086 | 0.06 ± 0.0074 |
| Myocardial infarction | Genetics | CAD PRS | 0.0401 ± 0.0035 | 0.0259 ± 0.0031 | 0.0626 ± 0.0073 |
| Myocardial infarction | Genetics | ISS PRS | 0.0339 ± 0.0041 | 0.0231 ± 0.0061 | 0.0549 ± 0.0106 |
| Myocardial infarction | Genetics | HT PRS | 0.0323 ± 0.0033 | 0.0215 ± 0.0065 | 0.051 ± 0.0057 |
